# First genotype-phenotype study in TBX4 syndrome: gain-of-function mutations causative for lung disease

**DOI:** 10.1101/2022.02.06.22270467

**Authors:** Matina Prapa, Mauro Lago-Docampo, Emilia M. Swietlik, David Montani, Mélanie Eyries, Marc Humbert, Carrie C.L. Welch, Wendy Chung, Rolf M.F. Berger, Ham Jan Bogaard, Olivier Danhaive, Pilar Escribano-Subías, Henning Gall, Barbara Girerd, Ignacio Hernandez-Gonzalez, Simon Holden, David Hunt, Samara M.A. Jansen, Wilhelmina Kerstjens-Frederikse, David Kiely, Pablo Lapunzina, John McDermott, Shahin Moledina, Joanna Pepke-Zaba, Gary J. Polwarth, Gwen Schotte, Jair Tenorio-Castaño, A.A. Roger Thompson, John Warton, Stephen J. Wort, NIHR BioResource for Translational Research – Rare Diseases, National Cohort Study of Idiopathic and Heritable PAH, PAH Biobank Enrolling Centers’ Investigators, Karyn Megy, Rutendo Mapeta, Carmen M. Treacy, Jennifer M Martin, Wei Li, Andrew J. Swift, Paul D. Upton, Nicholas W. Morrell, Stefan Gräf, Diana Valverde

## Abstract

**Rationale:** Despite the increasing frequency of *TBX4*-associated pulmonary arterial hypertension (PAH), genotype-phenotype associations are lacking and may provide important insights.

**Methods:** We assembled a multi-center cohort of 137 patients harboring monoallelic *TBX4* variants and assessed the pathogenicity of missense variation (n = 42) using a novel luciferase reporter assay containing T-BOX binding motifs. We sought genotype-phenotype correlations and undertook a comparative analysis with PAH patients with *BMPR2* causal variants (n = 162) or no identified variants in PAH-associated genes (n = 741) genotyped via the NIHR BioResource - Rare Diseases (NBR).

**Results:** Functional assessment of *TBX4* missense variants led to the novel finding of gain-of-function effects associated with older age at diagnosis of lung disease compared to loss-of-function (p = 0.038). Variants located in the T-BOX and nuclear localization domains were associated with earlier presentation (p = 0.005) and increased incidence of interstitial lung disease (p = 0.003). Event-free survival (death or transplantation) was shorter in the T-BOX group (p = 0.022) although age had a significant effect in the hazard model (p = 0.0461). Carriers of *TBX4* variants were diagnosed at a younger age (p < 0.001) and had worse baseline lung function (FEV1, FVC) (p = 0.009) compared to the *BMPR2* and no identified causal variant groups.

**Conclusions:** We demonstrated that TBX4 syndrome is not strictly the result of haploinsufficiency but can also be caused by gain-of-function. The pleiotropic effects of TBX4 in lung disease may be in part explained by the differential effect of pathogenic mutations located in critical protein domains.

## Introduction

Pulmonary Arterial Hypertension (PAH) is a rare disorder characterized by abnormal cell proliferation in the pulmonary arterioles, resulting in vascular occlusion and increased pulmonary artery pressure, ultimately leading to right ventricular failure (1). The introduction of high-throughput sequencing led to the discovery of several PAH-causal genes, although approximately 75% of idiopathic cases remain unexplained (2–4). Monoallelic pathogenic variants in the T-BOX transcription factor 4 (*TBX4*) gene are the second commonest heritable cause of PAH, often enriched in pediatric cohorts (5–7). TBX4 belongs to the T-BOX family of transcription factors playing a critical role in early hindlimb development (8–10) and branching of the lungs (11) regulating the expression of Fibroblast Growth Factor 10 (FGF10) together with TBX5 (12, 13). Transcriptome analysis suggests it continues to be active following organogenesis and plays an important role in the cellular homeostasis of adult lung fibroblasts (14).

*TBX4* sequence variants and contiguous gene deletions, as part of the recurrent chromosome 17q23.2 microdeletion, were originally reported in association with small patella syndrome (SPS) [MIM# 147891] (15–17). More recently, evidence is emerging that *TBX4* single nucleotide variants (SNVs)/deletions are not only causative of PAH, but also of a wide spectrum of developmental parenchymal lung disorders (18, 19). Partial or complete loss of a single *TBX4* functional allele is sufficient for the production of the phenotype, termed haploinsufficiency. Although the above disorders are dominantly inherited, both variable expressivity and reduced penetrance have been observed (5).

Herein, we assembled a large cohort of patients harboring *TBX4* sequence variants to establish genotype-phenotype correlations. We developed *in vitro* assays to assess the pathogenicity of non-truncating variants. This led to the novel finding of functionally distinguishable missense variation, causing either gain-(GoF) or loss-of-function (LoF).

## Methods

Using the term “TBX4”, our PubMed search identified 22 publications limited to human studies dating from 2004 to 2021 (supplementary data, xlsx). Cases with the recurrent 17q23.2 deletion were excluded from this study’s scope. We collected phenotypic information on 137 heterozygous carriers of *TBX4* sequence variants, including 15 novel cases from the National Institute for Health Research BioResource–Rare Diseases (NBR) study (20), the UK National Cohort Study of Idiopathic and Heritable PAH (2, 4), the Spanish registry of PAH (21), the registry of the French PAH network (22, 23), and the DECIPHER database (24). Demographic and phenotypic information at the time of diagnosis was captured from relevant publications and databases (supplementary data, xlsx). Follow-up information was obtained, where available.

Annotation of variants was harmonized to the *TBX4* canonical transcript NM_018488.3 of the human reference genome assembly GRCh37/hg19 using the Mutalyzer and Ensembl Variant Effect Predictor web services. We assessed variant pathogenicity according to the American College of Medical Genetics and Genomics (ACMG) guidelines using the VarSome Clinical tool followed by manual curation (25). Non-truncating variants (missense, indels) and variants predicted to affect splicing were selected for functional characterization (supplementary data, docx). Truncating variants (frameshift, nonsense) were presumed to cause LoF; p.Tyr127Ter pathogenic variant was used as a positive control in functional experiments.

Among the T-BOX family members, TBX4 has the highest similarity to TBX5 (26). Since no structure for TBX4 has been reported, we analyzed the effect of variants using the crystal structure of the TBX5 complex with DNA (Figure 3) (27). Variants were sub-grouped by protein domains, including the highly-conserved T-BOX domain (codons 71-251) containing the first nuclear localization segment NLS1 (91-103), and the predicted NLS2 (338-351) and transactivating region (−393) (28, 29).

We sought genotype-phenotype associations in carriers of likely pathogenic/pathogenic *TBX4* variants (excluded functionally assessed variants classed as benign/likely benign or of uncertain significance) and undertook a comparative analysis with genotyped PAH patients recruited to the NBR study, termed in this paper as BMRP2 (n = 162) and no identified causal variant (n = 741) groups (Table 1). The latter included patients with idiopathic PAH and likely pathogenic/pathogenic BMRP2 variants or no identified likely pathogenic/pathogenic variants in PAH associated genes (*BMPR2, TBX4, EIF2AK4, SMAD1/4/9, CAV1, KCNK3, ENG, ALK1, GDF2, AQP1, ATP13A3, SOX17*) (2). We additionally obtained computed tomography (CT) chest images from a subset of patients from the *BMPR2* (n = 34) and no identified causal variant (n = 143) groups and compared these to 11 *TBX4* cases from the NBR study. Details of the radiological sub-study can be found in the online supplement (docx).

**Table 1.** Demographic and clinical characteristics at diagnosis of the patient population included in the genotype-phenotype dataset.

|  | ***BMPR2***<br>**(n=162)** | **No causal**<br>**variant**<br>**identified**<br>**(n=741)** | ***TBX4***<br>**(n=98)** | **p value** | **Available**<br>**(total n)** |
| --- | --- | --- | --- | --- | --- |
| **Primary diagnosis** |  |  |  |  | 1001 |
| 1.1 Idiopathic PAH<br>*including drug- and toxin-<br>induced | 107 (66.0%) | 741<br>(100%) | 58<br>(59.2%) |  |  |
| 1.2 Heritable PAH | 53<br>(32.7%) | - | 17<br>(17.3%) |  |  |
| 1.4.1 PAH associated with<br>connective tissue disease | 1<br>(0.62%) | - | 2 (2.04%) |  |  |
| 1.4.4 PAH associated with<br>congenital heart disease | 1<br>(0.62%) | - | 11<br>(11.2%) |  |  |
| 1.6 PAH with overt<br>features of venous /<br>capillaries (PVOD/PCH)<br>involvement | - | - | 1<br>(1.02%) |  |  |
| 3.5 Developmental lung<br>disorders | - | - | 9<br>(9.18%) |  |  |
| **Sex: female** | 107 (66.0%) | 530<br>(71.5%) | 62<br>(63.9%) | 0.156 | 1000 |
| **Smoking history:**<br>**past/current** | 53<br>(39.0%) | 294<br>(51.8%) | 10<br>(30.3%) | 0.007 | 737 |
| **Exposure to drug or**<br>**toxins: yes** | 6<br>(3.70%) | 45<br>(6.07%) | 7<br>(19.4%) | 0.007 | 939 |
| **Ethnicity** |  |  |  |  | 966 |
| European | 137 (84.6%) | 630<br>(85.0%) | 53<br>(84.1%) |  |  |
| Finnish-European | - | 1 (0.13%) | - |  |  |
| African | 2<br>(1.23%) | 20<br>(2.70%) | 5<br>(7.94%) |  |  |
| East-Asian | 2<br>(1.23%) | 6<br>(0.81%) | 1<br>(1.59%) |  |  |
| South-Asian | 6<br>(3.70%) | 48<br>(6.48%) | 1<br>(1.59%) |  |  |
| Other | 15 (9.26%) | 36 (4.86%) | 3 (4.76%) |  |  |
| **Age at diagnosis of lung disease (years)** | 39 [31;51] | 51 [38;66] | 14 [2;48] | <0.001 | 997 |
| **Age at transplantation or death (years)** | 52 [43;61] | 67 [53;75] | 64 [1;71] | <0.001 | 302 |
| **WHO functional class** |  |  |  | 0.277 | 918 |
| I | 2<br>(1.24%) | 15<br>(2.11%) | 2<br>(4.44%) |  |  |
| II | 32<br>(19.9%) | 144<br>(20.2%) | 10<br>(22.2%) |  |  |
| III | 96<br>(59.6%) | 466<br>(65.4%) | 27<br>(60.0%) |  |  |
| IV | 31<br>(19.3%) | 87<br>(12.2%) | 6<br>(13.3%) |  |  |
| **Exercise test** |  |  |  |  |  |
| Distance (meters) | 350<br>[276;420] | 330 [210;410] | 371<br>[308;422] | 0.028 | 810 |
| Pre-test sats | 96.0<br>[94.0;98.0] | 96.0<br>[93.0;97.0] | 97.5<br>[95.5;98.0] | 0.006 | 738 |
| Post-test sats | 94.0<br>[89.0;96.8] | 91.0<br>[85.0;95.0] | 95.5<br>[86.5;97.0] | 0.001 | 683 |
| **Lung function (%pred)** |  |  |  |  |  |
| FEV1 | 91.0<br>[79.0;100] | 85.0<br>[72.3;96.0] | 82.0<br>[70.0;98.0] | 0.001 | 719 |
| FVC | 99.7 (17.2) | 93.2 (19.4) | 88.0 (17.7) | <0.001 | 703 |
| KCO | 83.4<br>[74.2;96.5] | 68.0<br>[49.0;83.0] | 72.0<br>[59.8;89.8] | <0.001 | 516 |
| TLC | 96.0<br>[89.0;106] | 94.0<br>[85.0;104] | 104<br>[98.0;110] | 0.044 | 511 |
| **Hemodynamics** |  |  |  |  |  |
| mPAP(mmHg) | 57.0<br>[52.0;66.8] | 52.0<br>[42.0;61.0] | 60.5<br>[48.2;82.2] | <0.001 | 916 |
| mPAWP(mmHg) | 10.0<br>[7.00;12.0] | 9.00<br>[7.00;12.0] | 9.00<br>[7.00;11.0] | 0.841 | 822 |
| PVR(WU) | 14.5<br>[10.8;20.4] | 10.3<br>[7.06;13.9] | 12.8<br>[8.25;16.2] | <0.001 | 767 |
| CO(L/min) | 3.30<br>[2.69;3.94] | 4.04<br>[3.25;5.10] | 3.65<br>[3.09;4.61] | <0.001 | 852 |
| CI(L/min/m<sup>2</sup>) | 1.90<br>[1.51;2.23] | 2.30<br>[1.80;2.80] | 2.63<br>[1.98;3.20] | <0.001 | 503 |
| Vasoresponders | 1 (1.28%) | 51 (17.7%) | 6 (10.2%) | 0.001 | 425 |
| **Increased BNP (>50**<br>**pg·mL<sup>-1</sup>)**<br>**or NT-proBNP (>300**<br>**pg·mL<sup>-1</sup>)** | 34 (97.1%) | 140 (79.1%) | 5 (71.4%) | 0.011 | 219 |
| Abbreviations: % pred, percentage of predicted value; BNP, brain natriuretic peptide; CI, cardiac index; CO, cardiac output; FEV1; forced expiratory volume in one second; FVC, forced vital capacity; mPAP, mean pulmonary artery pressure; KCO, carbon monoxide transfer coefficient; mPAWP, mean pulmonary artery wedge pressure; NT-pro-BNP, N-terminal pro-brain natriuretic peptide; PCH, pulmonary capillary hemangiomatosis; PVOD, pulmonary veno-occlusive disease; PVR, pulmonary vascular resistance; TLC, total lung capacity; WHO, World Health Organization. |  |  |  |  |  |

Statistical analysis was performed using R (www.r-project.org). The R package “survival” was used to compare event-free survival between different groups. Survival was estimated by the Kaplan-Meier method from the time of diagnosis to death or transplantation. To avoid immortal time bias, this was limited to a 10-year interval. Gender and age at diagnosis of lung disease were included as covariates in the semi-parametric Cox-proportional hazard models.

## Results

### Study population

We identified 137 heterozygous carriers of *TBX4* variants, the majority of which were sporadic cases (n=127, 93%). Out of four identified families, eight related individuals with available detailed phenotypic information were included in our analyses. Twenty-one cases had a primary phenotype of SPS. In the remaining 116 individuals presenting with lung disease, subsequent assessment for SPS was lacking in most, with only 29 cases (25%) reported having associated skeletal features. PAH was the predominant primary lung phenotype (Table 1). Median age at diagnosis of lung disease was 14 years (IQR: 2-47 years). Fifty-three individuals (45%) presented in adulthood, 41 (36%) in childhood, and 22 (19%) in the perinatal period. In the overall patient cohort, there was an equal female-to-male ratio with an observed female predominance (62%) in the lung disease group.

### Spectrum and functional assessment of *TBX4* variants

A total of 108 distinct *TBX4* variants were retrieved from the literature and aforementioned databases (Figure 1). Of these, 43 were missense, 39 frameshift, 15 nonsense, 6 indels including an additional deletion of whole exon 5, and 3 variants predicted to affect splicing (supplementary data, xlsx). A single case with a variant in the *TBX4* promoter region was excluded from our genotype-phenotype analyses (22).

**Figure 1.**
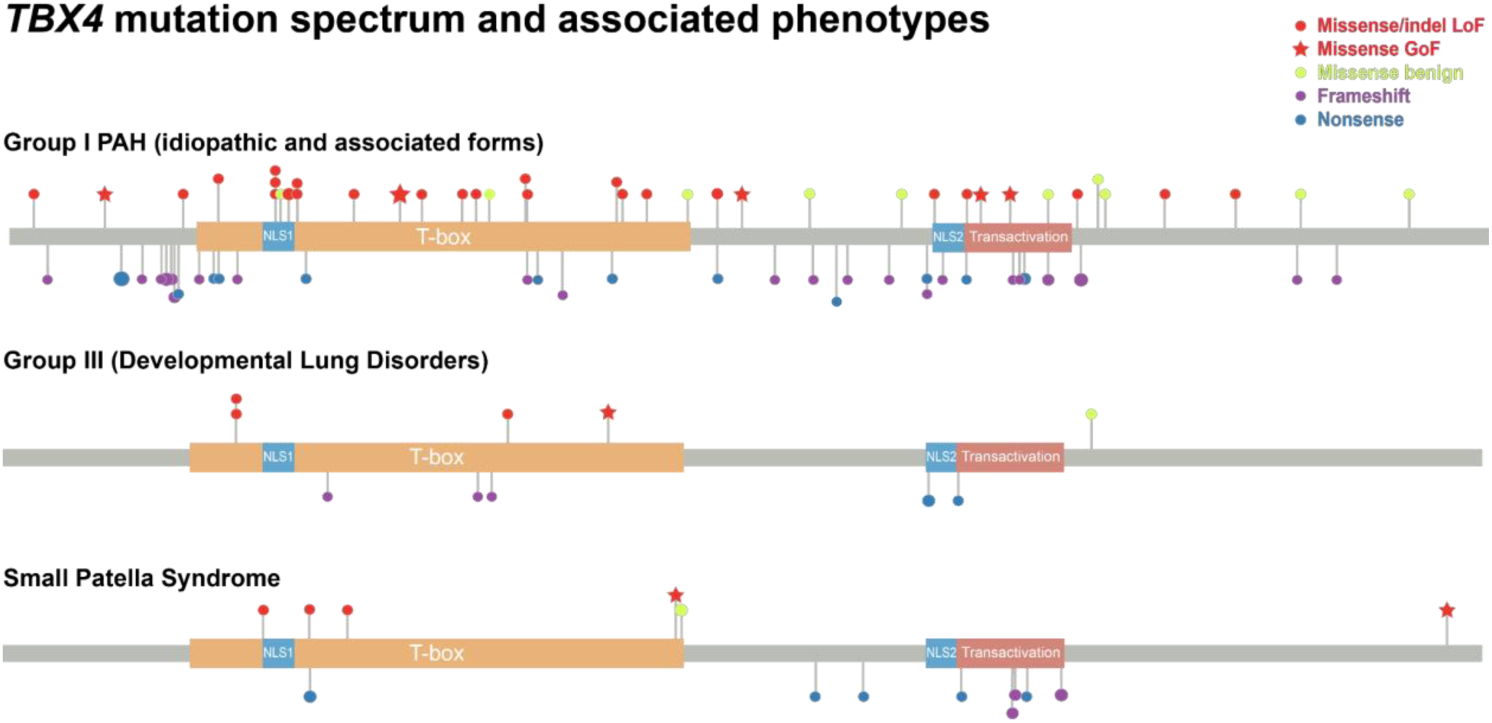
Lollipop plot depicting *TBX4* mutation spectrum. Recurrently mutated positions are represented by a proportionally sized lollipop. Critical protein domains are highlighted, inclusive of the DNA binding T-BOX, nuclear localization segments (NLS1 and NLS2), and transactivation domains. Variants are grouped by primary associated phenotype and color-coded taking into account the functional assessment of missense and inframe insertion/deletions (indels). Abbreviations: GoF, gain-of-function; LoF, loss-of-function.

We assessed the pathogenicity of all indels and 42/43 missense variants using a luciferase reporter assay (Figure 2). Variant c.1021G>C was assessed by a minigene assay instead as predicted to affect the same donor splice site as in c.1021+1G>A, which resulted in a double exon skipping. All indels and 23 missense variants caused LoF with another 11 shown to be benign. Eight missense variants resulted in GoF with a mean Relative Luciferase Units (RLU) of approximately twice the levels of the wild-type construct. We confirmed that GoF was not an artifact by checking TBX4 protein expression levels of several constructs with different outcomes in the luciferase assay by qPCR and western-blot (supplementary eFigure 1). Independent of their functional activity, all assessed variants translocated to the nucleus, replicating the translocation of wild-type and Green Fluorescent Protein tagged TBX4 (supplementary eFigure 2 & 3).

**Figure 2.**
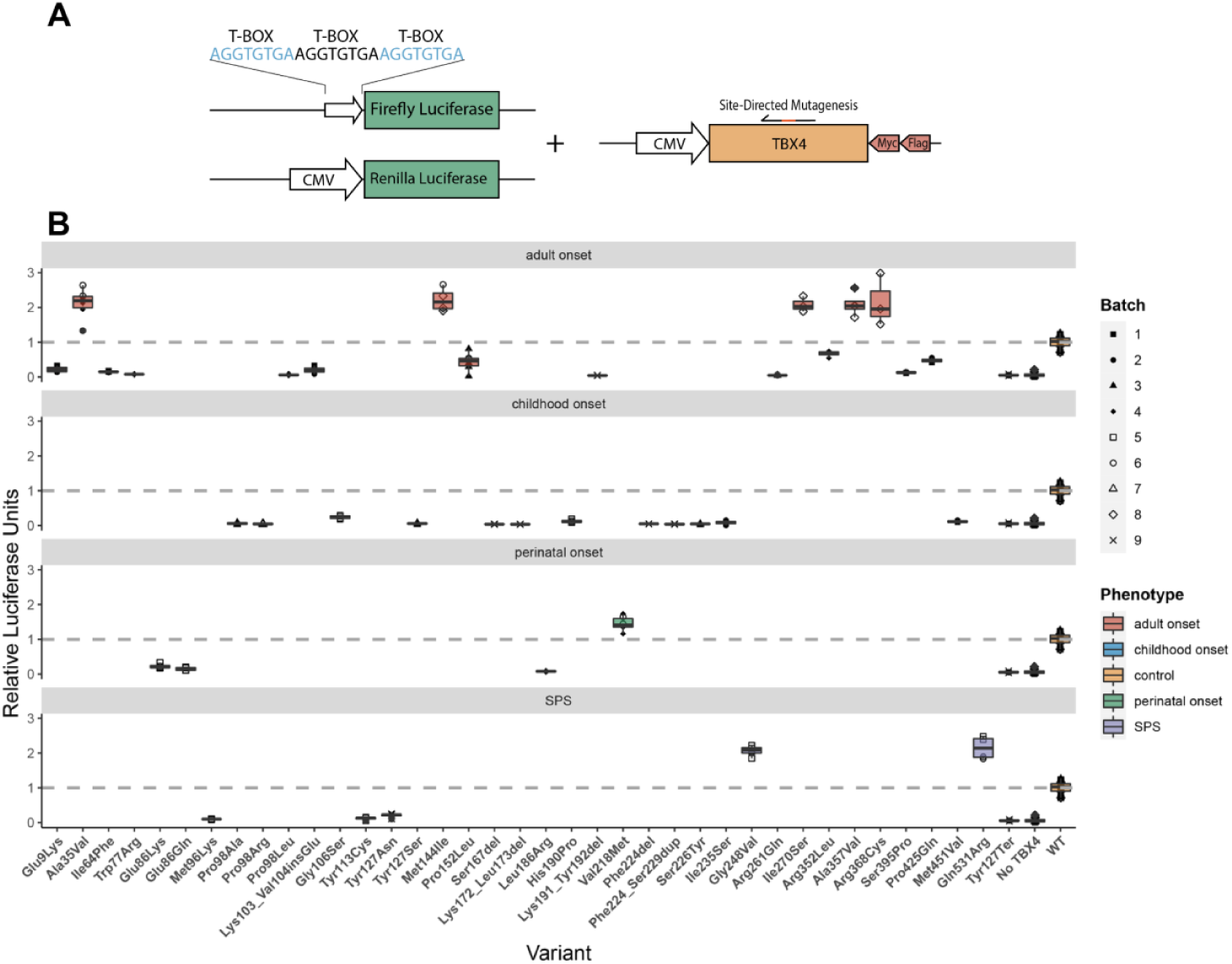
Functionally assessed *TBX4* variants by luciferase assay inducing gain- or loss-of-function. A) Schematic representation of the *in vitro* dual-luciferase reporter assay. We co-transfected three different plasmids: Firefly luciferase with x3 T-BOX motifs as the promoter, Renilla luciferase, and TBX4 overexpression plasmid (wild type/mutated). B) Variants inducing gain- or loss-of-function grouped by primary phenotype; lung disease (perinatal-, childhood-, and adult-onset) or small patella syndrome (SPS). The y-axis represents the Relative Luciferase Units; the dashed line marks the level of the wild type. Data is shown as box plots representing median ± quartiles. Dots represent biological replicates with corresponding batches in different shapes.

**Figure 3.**
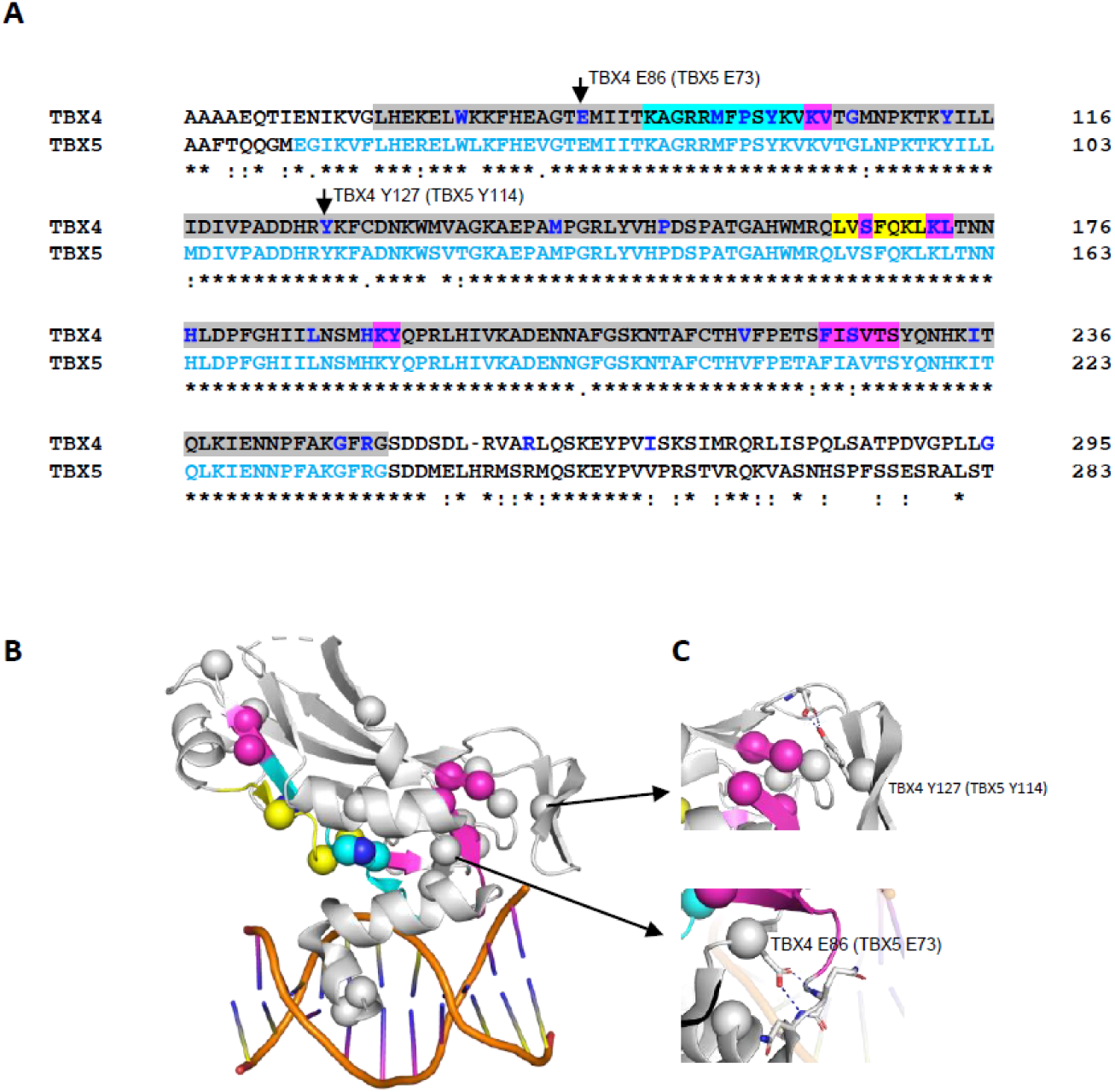
Structural analysis of TBX4 sequence variants. The crystal structure of TBX5 bound to DNA, pdb code 2×6V, was used for structural analysis. They share 52.6% sequence along with the full-length proteins and 93.9% in the T-BOX domain. A) Sequence alignment of TBX4 and TBX5 in the T-BOX region (highlighted in grey) containing the DNA-binding motif as well as the nuclear localization segment 1 (NLS1, in cyan) and the nuclear export segment (NES, in yellow)(28, 29). TBX4 missense variants are indicated in bold/blue, with indels highlighted in magenta. Residues visible in the TBX5 structure are shown in light blue letters. B) Mutations plotted on the TBX5 crystal structure as spheres. Cyan, yellow, and magenta spheres correspond to the NLS1, NES regions, and indels as indicated in A. When annotating loss-of-function variants on the TBX4 sequence, they are highly enriched in the T-BOX, particularly the NLS1 and NES. C) Some mutations of the non-interface residues, such as TBX4 p.Glu86 and p.Tyr127 (corresponding residues p.Glu73 and p.Tyr114 in TBX5, respectively), make essential interactions to stabilize the secondary structural elements required for T-BOX binding to DNA. Clustal Omega was used for sequence alignment. Figures were generated using PyMOL Molecular Graphics System.

As per ACMG guidelines, our luciferase functional data altered the classification of the majority of respective variants with a total of 33/48 (67%) initially classified as variants of uncertain significance, and an equivalent number (32/48) of likely pathogenic/pathogenic variants following application of the PS3 criterion (*in vitro* functional studies supportive of a damaging effect) where appropriate (Supplementary eFigure 4). We evaluated the performance of *in silico* tool predictions for likely pathogenic/pathogenic *TBX4* missense variants, with SIFT generating the highest overall percentage of correct calls (Supplementary eFigure 5). Overall, a total of 4/8 GoF and 22/23 LoF missense variants were classed as likely pathogenic/pathogenic and included in the genotype-phenotype analyses (supplementary data, xlsx).

**Figure 4.**
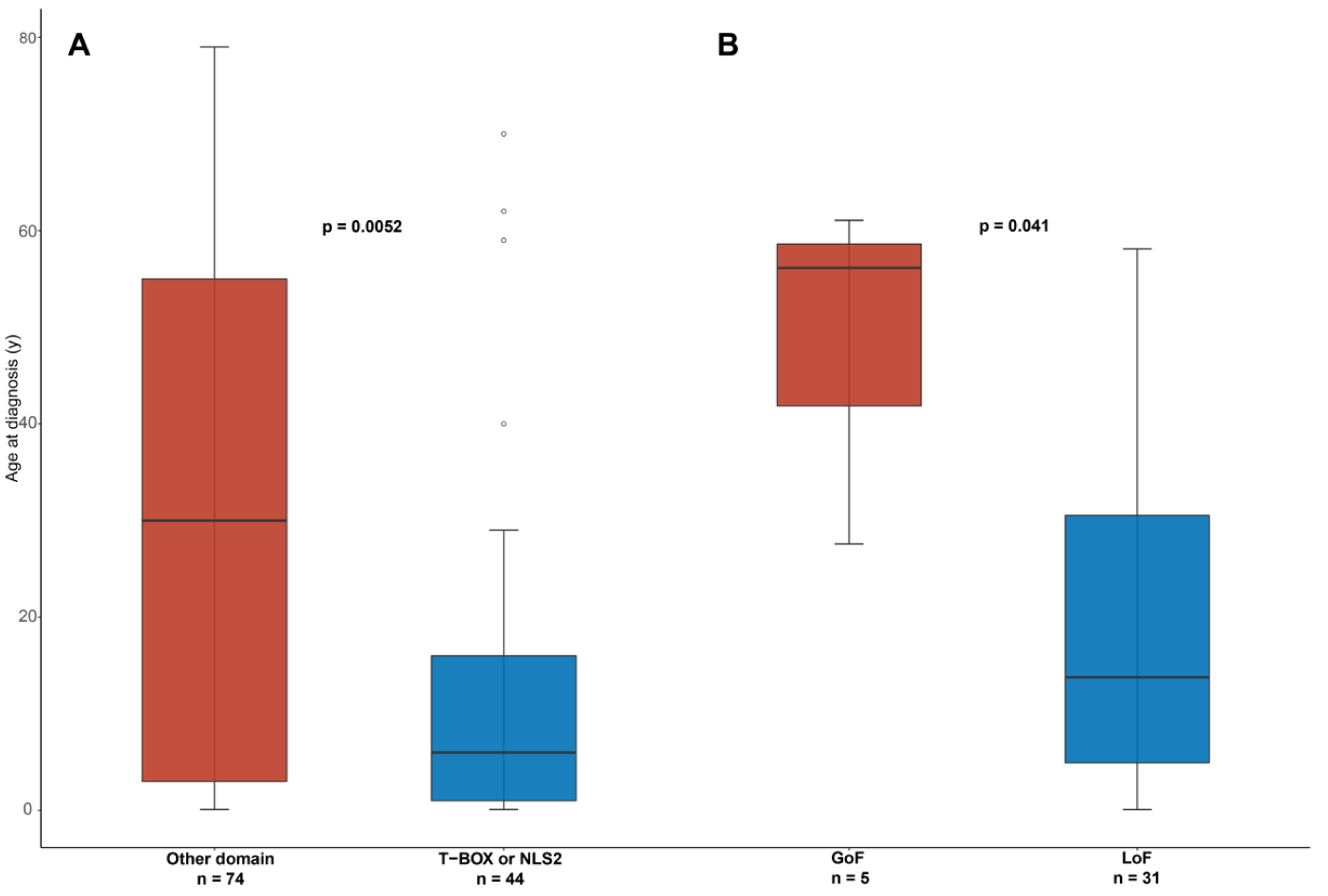
Age at diagnosis of lung disease by *TBX4* genotype; (A) variants in the T-BOX and second nuclear localization segment (NLS2) versus other domains, (B) likely pathogenic/pathogenic gain-of-function versus loss-of-function missense variants.

**Figure 5.**
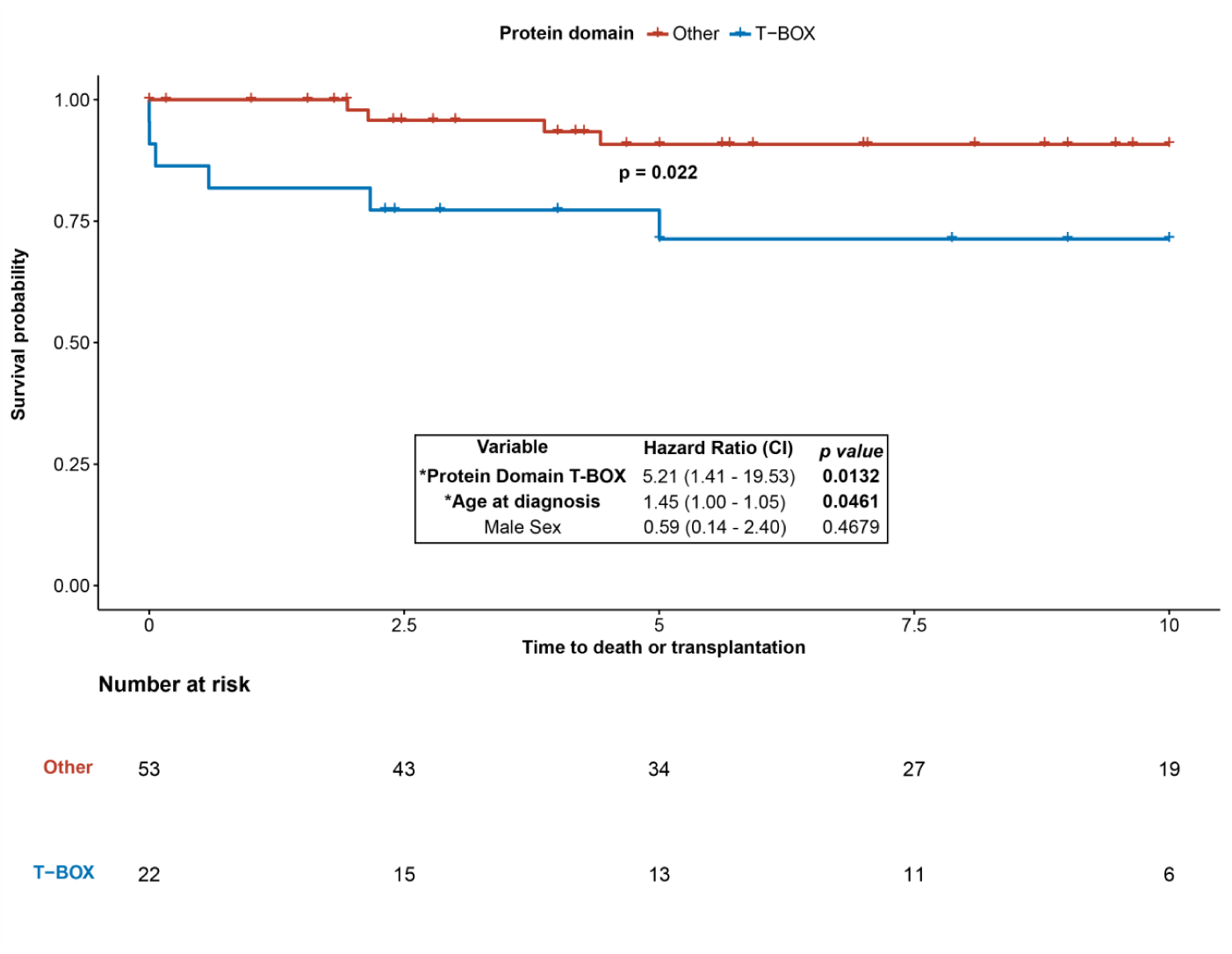
Time to death or lung transplantation (years) by TBX4 protein domain genotype. Event-free survival was shorter in the T-BOX domain variant group although age had a significant effect in the hazard model.

Finally, we assessed the functional impact of all previously reported *TBX4* variants predicted to affect splicing with recurrent variants c.702+1G>A and c.1021+1G>A inducing exon skipping events (Supplementary eFigure6). Key findings of the structural variant analysis are summarized in Figure 3.

### Genotype-phenotype associations

Patients with lung disease and variants within the DNA binding T-BOX domain presented at a younger age (median [IQR]: 7.5 [1 - 18.5] years) compared to carriers of variants outside this domain (18 [3 - 51.5] years, p = 0.028). This remained true for sequence variants located in either the T-BOX domain containing NLS1 or NLS2 at the C-terminus (p = 0.005, Figure 4). Individuals with LoF variants (missense, indels) were also diagnosed at a younger age compared to GoF variant carriers (14 [5 - 31] years vs. 57 [42.5 - 59.5], p = 0.038, Figure 4).

Baseline clinical features and hemodynamic parameters did not differ significantly between protein domain groups with the exception of interstitial lung disease which was more frequently reported in carriers of *TBX4* variants located in the T-BOX domain alone (75% vs. 21%, p = 0.003) or in combination with NLS2 variants (p = 0.001). Similarly, no significant differences were observed between LoF and GoF variation and this was consistent when protein-truncating variants were added to the LoF group.

The observed primary phenotypes varied significantly between protein domains with a greater frequency of developmental lung disorders (including acinar dysplasia and congenital alveolar dysplasia) in the T-BOX group (15.4% vs. 3.8%, overall p = 0.046); when combined with variants in the NLS2 domain, this remained statistically significant (overall p = 0.003). Lung histology was available in 17 previously published *TBX4* cases (see supplementary docx and xlsx for details). A greater frequency of a confirmed secondary phenotype of SPS was observed in carriers of variants located outside the T-BOX and NLS2 domains (29.7% vs. 11.4%, p = 0.038) and was more prevalent in protein-truncating versus missense variation (27.8% vs. 7.1%, p = 0.044). Variant exon location did not appear to have any phenotypic impact (data not shown).

As previously reported (30), patients with TBX4 associated lung disease presented at a younger age (median [IQR]: 14 [2 - 48] years) compared to patients in the *BMPR2* (39 [31 – 51] years) and no identified causal variant (51 [38 - 66] years, p < 0.001) groups. They performed better at the six-minute walk test with no significant differences in functional class at presentation (Table 1). They also had worse baseline lung function (FEV1, FVC) despite presentation at a younger age (Table 1). The frequency of airway / acinar abnormalities was significantly greater in the *TBX4* (87.5%) versus the *BMPR2* (32.4%) and no identified causal variant groups (33.6%, p = 0.009) (Table 2).

**Table 2.** Radiological features of Computed Tomography (CT) of the chest in pulmonary arterial hypertension (PAH) cases analyzed as part of the imaging sub-study.

|  | ***BMPR2***<br>**(n=34)** | **No causal**<br>**variant identified**<br>**(n=143)** | ***TBX4***<br>**(n=8)** | **p**<br>**value** | **Available**<br>**(total n)** |
| --- | --- | --- | --- | --- | --- |
| **Vascular features:** |  |  |  |  |  |
| **Axial MPA diameter**<br>**(mm)** | 34.2<br>[30.9;37.4] | 34.6 [31.4;38.3] | 34.0<br>[30.7;38.5] | 0.674 | 185 |
| **PA to Ao ratio** | 1.23<br>[1.12;1.40] | 1.14 [0.99;1.29] | 1.18<br>[0.94;1.32] | 0.083 | 185 |
| **Neovascularity** | 7 (23.3%) | 10 (7.69%) | - | 0.048 | 167 |
| **Lung & mediastinal**<br>**features:** |  |  |  |  |  |
| **Ground-glass**<br>**opacities** | 18 (52.9%) | 62 (43.4%) | - | 0.016 | 185 |
| *Centrilobular pattern |  |  |  | 0.088 | 185 |
| None | 17 (50.0%) | 90 (62.9%) | 8 (100%) |  |  |
| Subtle | 4 (11.8%) | 21 (14.7%) | - |  |  |
| Present | 13 (38.2%) | 32 (22.4%) | - |  |  |
| **Non-specific mosaic<br>pattern |  |  |  | 0.441 | 180 |
| None | 31 (91.2%) | 130 (90.9%) | 3 (100%) |  |  |
| Subtle | - | 7 (4.90%) | - |  |  |
| Present | 3 (8.82%) | 6 (4.20%) | - |  |  |
| **Fibrosis** |  |  |  | 1.000 | 185 |
| None | 34 (100%) | 137 (95.8%) | 8 (100%) |  |  |
| Present | - | 6 (4.2%) | - |  |  |
| **Pleural effusion** |  |  |  |  |  |
| None | 33 (97.1%) | 132 (92.3%) | 8 (100%) | 1.000 | 185 |
| Subtle | - | 3 (2.10%) | - |  |  |
| Present | 1 (2.94%) | 8 (5.59%) | - |  |  |
| **Interlobular septal thickening** |  |  |  | 0.157 | 179 |
| None | 27 (87.1%) | 122 (87.1%) | 5 (62.5%) |  |  |
| Subtle | 3 (9.68%) | 9 (6.43%) | 1 (12.5%) |  |  |
| Present | 1 (3.23%) | 9 (6.43%) | 2 (25.0%) |  |  |
| **Mediastinal lymphadenopathy** |  |  |  | 0.721 | 185 |
| None | 29 (85.3%) | 122 (85.3%) | 6 (75.0%) |  |  |
| Present | 5 (14.7%) | 21 (14.7%) | 2 (25.0%) |  |  |
| **Airway/acinar features:** | 11 (32.4%) | 48 (33.6%) | 7 (87.5%) | 0.009 | 185 |
| **Bronchial wall thickening** |  |  |  | 0.051 | 179 |
| None | 26 (83.9%) | 126 (90.0%) | 6 (75.0%) |  |  |
| Subtle | 3 (9.68%) | 11 (7.86%) | - |  |  |
| Present | 2 (6.45%) | 3 (2.14%) | 2 (25.0%) |  |  |
| **Emphysema** |  |  |  | 0.026 | 185 |
| None | 32 (94.1%) | 120 (83.9%) | 5 (62.5%) |  |  |
| Subtle | - | 13 (9.09%) | - |  |  |
| Present | 2 (5.88%) | 10 (6.99%) | 3 (37.5%) |  |  |
| **Air trapping** |  |  |  | 0.040 | 185 |
| None | 28 (82.4%) | 121 (84.6%) | 4 (50.0%) |  |  |
| Subtle | 4 (11.8%) | 7 (4.90%) | 1 (12.5%) |  |  |
| Present | 2 (5.88%) | 15 (10.5%) | 3 (37.5%) |  |  |
| **Suspected PVOD:** | 1 (3.03%) | 8 (5.59%) | 1 (12.5%) | 0.500 | 184 |
| Abbreviations: Ao, aortic; MPA, main pulmonary artery; PA, pulmonary artery; PVOD, pulmonary veno-occlusive disease. |  |  |  |  |  |

### Impact of genotype on survival

Clinical outcomes were available for 89/115 *TBX4* cases presenting with lung disease with a median follow-up of 8 years (IQR: 3 - 10 years). Event-free survival was shorter in the T-BOX domain variant group (p = 0.022 for log-rank test) although age at diagnosis also had a significant effect (p = 0.0461 for Cox-proportional hazard model, Figure 5). There were no observed differences in outcomes between GoF and LoF variants or missense versus protein-truncating variants. Overall, event-free survival was longer in the *TBX4* group compared to *BMPR2* and no identified causal variants groups (p = 0.0025). Pairwise comparisons showed no significant differences between *TBX4* and *BMPR2* variant carriers (p = 0.69). Compared to individuals with no causal sequence variants, both patients with *TBX4* (p = 0.035) and *BMPR2* variants (p = 0.016) had higher event-free survival rates. However, genotype did not have a significant effect on survival following correction for age and sex; both male gender (p = 0.0002) and older age at the time of diagnosis (p < 2e-16) were associated with shorter survival in the Cox-proportional hazard model.

## Discussion

### Distinct mutational mechanisms disrupt TBX4 function

A wide range of *TBX4* mutations can result in human disease. Intragenic sequence variants occur throughout the gene and are mostly private. The functional impact of *TBX4* variants was not previously investigated and a haploinsufficient effect was assumed. Our assessment of *TBX4* missense variants reported to date led to the discovery of two distinct functional classes of variants, GoF or LoF. Although this constitutes a novel finding, GoF variants have been reported in other members of the T-BOX gene family in association with a similar phenotypic spectrum caused by LoF. This includes carriers of missense variants in the TBX1 gene with velocardiofacial syndrome, a single TBX5 variant in a family with Holt-Oram syndrome, and a TBX20 variant in a family with atrial septal defects and valvular disease (31–33). All of the above were located in the highly-conserved T-BOX domain with the exception of a single TBX1 variant in exon 8 of 9 (c.928G>A). Structural analyses were suggestive of increased protein stability; our respective analysis was indicative of the detrimental effects of LoF variants with no simple explanation for GoF variation (Figure 3).

Out of 31 pathogenic missense *TBX4* variants reported in our study, 8 resulted in GoF including the recurrent variant c.432G>T (21). These were located across the gene with 3 in the T-BOX region and 2 in the transactivation domain (Figure 1). Two variants (c.743G>T and c.1592A>G) were originally reported in 2004 in 2 Dutch families with classical SPS and constituted one of the first reports implicating *TBX4* in this phenotype (15). The remaining ones were described in association with adult-onset lung disease diagnosed as late as 81 years with the exception of a neonate presenting with interstitial lung disease (Table 3). Notably, 6/8 GoF variants were present in control populations (gnomAD database) including variant c.104C>T with the highest overall population frequency in the cumulative *TBX4* variant dataset. As a result, many of these were classified as likely benign/uncertain clinical significance and cannot be considered as responsible for the underlying phenotype at present (Table 3).

**Table 3.** Clinical features of individuals heterozygous for *TBX4* gain-of-function missense variants

| Variant | Demographics | Primary lung phenotype & age at diagnosis | Details of lung disease & co-morbidities | Skeletal features | Follow-up | Pedigree information | ACMG classification & population frequency (gnomAD exomes) |
| --- | --- | --- | --- | --- | --- | --- | --- |
| c.104C>T;<br>p.Ala35Val | *Dutch PAH patient cohort* (39) | PAH adult-onset | N/A | N/A | Deceased (unavailable details) | N/A | Likely Benign<br>0.009 |
| c.432G>T;<br>p.Met144Ile | Female, White Hispanic (21) | IPAH 28 years | Clinical diagnosis of possible PVOD due to radiological findings and reduced diffusion capacity. Histopathological findings (explanted lung tissue): typical PAH features, no evidence of PVOD or ILD | Absent | Clinical deterioration with bilateral lung transplantation 9 years following diagnosis | Maternally inherited variant, PAH ruled out at 67 years | Pathogenic<br>3.98E-06 |
| c.432G>T;<br>p.Met144Ile | Male, White Hispanic (21) | PVOD 62 years | Radiological findings typical of PVOD. Heart failure signs, severe respiratory insufficiency. Not eligible for lung transplant due to advanced age and comorbidities | Absent | Deceased (26 months following diagnosis) | No known family history of PAH/SPS. Deceased parents prior to PAH diagnosis. Genetic counseling provided to the rest of the family (declined genetic testing) | Pathogenic<br>3.98E-06 |
| c.652G>A;<br>p.Val218Met | Female, White European (23) | chILD 5 months | Lung biopsy at 7 years: diffuse alveolar simplification, moderate thickened PA muscular wall, and PA fibrointimal proliferation. PFO, microcephaly, hearing loss | Short stature, no SPS features | 10 years: chILD, moderate pulmonary hypertension | Parents not tested | VUS<br>1.23E-04 |
| c.743G>T;<br>p.Gly248Val | Dutch family (15, 39) | - | Family members screened for PAH by echocardiography with none fulfilling diagnostic criteria (unavailable details) | Classical SPS phenotype, including patellar and pelvic anomalies | N/A | 3-generation pedigree (family A) (15) | Pathogenic<br>Not Reported |
| c.809T>G;<br>p.Ile270Ser | Female, White<br>European (36) | IPAH<br>57 years | Functional class III at diagnosis.<br>Asthma, HTN, obesity, sleep apnoea | N/A | Alive (1-year follow-up) | N/A | Likely Pathogenic<br>4.00E-06 |
| c.1070C>T;<br>p.Ala357Val | Female, White<br>British (2) | IPAH<br>81 years | Functional class III at diagnosis.<br>Radiological findings included minor<br>bronchial wall thickening, no<br>evidence of PVOD except<br>mediastinal nodes, no interstitial lung<br>disease. Hypothyroidism, IHD,<br>T2DM, HTN, breast cancer | N/A | Died in hospice 7<br>months following<br>diagnosis<br>(progressive heart<br>failure) | N/A | VUS<br>1.99E-05 |
| c.1102C>T;<br>p.Arg368Cys | Female, African-Caribbean (2) | IPAH<br>39 years | Functional class III at diagnosis.<br>Radiological findings are typical of<br>PAH, no lung disease, PFO,<br>hypothyroidism, chronic subdural<br>hematoma | N/A | Deceased<br>(50 years) | N/A | VUS<br>5.58E-05 |
| c.1592A>G;<br>p.Gln531Arg | Dutch family<br>(15, 39) | - | Family members screened for PAH<br>by echocardiography with none<br>fulfilling diagnostic criteria<br>(unavailable details) | Classical SPS<br>phenotype,<br>including<br>patellar and<br>pelvic<br>anomalies | N/A | 3-generation pedigree<br>(family C) (15) | Likely Pathogenic<br>Not Reported |
| Abbreviations: ACMG, American College of Medical Genetics and Genomics; chILD, childhood-onset interstitial lung disease; HTN, systemic hypertension; ILD, interstitial lung disease; IHD, ischaemic heart disease; IPAH, idiopathic pulmonary arterial hypertension; N/A, not available; PA, pulmonary artery; PAH, pulmonary arterial hypertension; PFO, patent foramen ovale; PVOD, pulmonary veno-occlusive disease; SPS, small patella syndrome; T2DM, Type 2 diabetes mellitus; VUS, variant of uncertain clinical significance. |  |  |  |  |  |  |  |

It remains to be seen whether these GoF variants are purely hypermorphic, increasing the protein’s function, or neomorphic, causing ectopic expression or acquisition of a new function. This may in turn influence their exerted phenotypic effects. From a mechanistic perspective, our knowledge of the consequences of TBX4 overexpression during embryogenesis and/or following organogenesis is lacking. Transcription can be affected not only by decreased but also by excessive amounts of transcription factors (34). Comparison of likely pathogenic/pathogenic gain- and loss-of-function missense variation showed a later onset lung disease in the former group (Figure 4), which may suggest a milder influence on the phenotype with additional genetic modifiers likely at play (19). A number of functionally assessed *TBX4* missense variants appeared to be hypomorphic, resulting in reduced levels of transcriptional activity but not complete LoF (Figure 2, eFigure 7); the vast majority of null variants were located in the T-BOX domain whereas hypomorphic variants spread across the gene. However, the above differences in transcriptional activity alone were not sufficient to explain the diverse phenotypic spectrum of TBX4 disease (no significant genotype-phenotype correlations).

### Modifiers of TBX4 disease spectrum

Our study captures the variable expressivity of *TBX4* pathogenic variation, with additionally observed phenotypic differences in recurring variants (supplementary data, xlsx). Aside from hypertensive pulmonary vascular disease, distal lung development can be disrupted to a variable degree raising the issue of TBX4 disease classification under World Health Organization Group 3, pulmonary hypertension associated with hypoxia and lung disease, as opposed to Group 1, idiopathic/heritable PAH. Although the underlying mechanisms are not yet fully elucidated, the impact of genotype was discernible with a greater frequency of developmental lung disorders and interstitial lung disease in individuals harboring pathogenic variants located in the T-BOX domain and nuclear localization segments, NLS1 and NLS2. In contrast, SPS was more frequently observed as a secondary phenotype when variants occurred outside the above TBX4 regions.

Despite the younger age of onset of lung disease in *TBX4* compared to *BMPR2* variant carriers, baseline lung function was worse with evidence of possible disrupted alveolarization on CT imaging differentiating this form of PAH as a primary lung developmental disorder. It can be postulated that this partly accounts for the variable age of onset of *TBX4* associated pulmonary hypertension with less pulmonary reserve and increased susceptibility to external insults acting as environmental modifiers of penetrance.

### Gene-specific variant classification and prognostic value

Inconsistent variant interpretation can not only lead to misdiagnosis of individual patients but also have significant consequences for at-risk relatives through inappropriate predictive testing. Gene-specific knowledge overcomes some of these pitfalls, especially when semi-automated impact analysis tools are used for variant classification. *In silico* predictions did not reliably reflect the true effects of *TBX4* missense variants on gene activity (supplementary eFigure 4). Structural analysis was also not capable of discerning *TBX4* GoF variants. In light of these limitations, an integrated pipeline incorporating molecular testing and functional assessment of novel *TBX4* missense variants by standardized assays would be of high diagnostic value.

Our results indicate that both protein-truncating and missense variants contribute to TBX4 disease with the majority of the latter (26/42) classified as likely pathogenic/pathogenic following functional assessment. In light of this, we removed the ACMG BP1 criterion (missense variant in a gene for which primarily truncating variants are known to cause disease) from *TBX4* variant annotation. In addition, the PM1 criterion (variant located in critical/well-established functional domain) could be expanded to include not only variants located in the T-BOX domain but also the predicted nuclear localization segments (NLS1 and NLS2) as our study was suggestive of an equivalent effect on produced phenotypes, including a higher rate of developmental (early-onset) lung disorders.

Estimating the true penetrance of TBX4 disease remains a high priority and impacts on variant interpretation as well as genetic counseling of respective families. A characteristic example exhibiting inter- and intra-familial variability is that of splice variant c.1021+1G>A reported by 3 independent studies (supplementary data, xlsx); index cases had adult-onset lung disease with positive family history, including first-degree relatives with severe PAH resulting in death in infancy/childhood or absent patella only (21, 35, 36). Leaky splicing variants (reducing but not completely abolishing the production of normal transcripts) can result in reduced penetrance although this phenomenon would still not explain the variable phenotypes observed in association with other types of recurrent variants (supplementary data, xlsx). We applied the ACMG BS1 criterion (allele frequency is greater than expected for the disorder) using a maximum tolerated population frequency of 5.00 × 10-8 arising from a generous estimate of 50% for lung disease penetrance (37); out of 17/42 missense *TBX4* variants present in gnomAD, several GoF and LoF variants remained of uncertain clinical significance (online supplement, docx and xlsx). Where there is enough evidence to support (likely) pathogenicity for missense variation in critical protein domains, important prognostic information can be inferred based on our genotype-phenotype analysis.

### Limitations

We were limited by the multi-center nature of this study including retrospective data collection with variable follow-up duration. We were unable to account for missing phenotypic information (presence/absence of SPS features in individuals with a primary lung phenotype), although estimation of TBX4 disease-penetrance was not the focus of this work. The main limitation of our functional assay is that we used overexpression plasmids, forcing the expression of variants whose effect could be lowered by post-transcriptional regulation. Our reporter system was designed using the standard T-BOX motif and FGF10 promoter sequences. A recent report of TBX4 chromatin immunoprecipitation in human fetal lung fibroblasts identified several other potential genome-wide target sites whose effects we have not tested (38). Therefore, a variant shown to have a modest or benign effect by our luciferase assay may still have significant damaging effects on interactions with other crucial binding sites in biologically relevant tissues.

## Conclusion

In summary, we combined functional and phenotypic characterization of all *TBX4* monoallelic variants published in the literature to date to determine the hallmarks of *TBX4*-mediated lung disease. We used *in vitro* analyses to assess the pathogenicity of missense, indel, and splice variants, resulting in either loss- or gain-of-function effects with phenotypic and prognostic implications when also taking into account variant location in functional domains. Our knowledge of *TBX4* genotype-phenotype associations can only be furthered by active collaborations between molecular scientists and clinicians, requiring both an indepth understanding of the biological aspects of the disease and a systematic approach to phenotyping.

## Supporting information

supplementary data, docx

supplementary data, xls

## Data Availability

All data produced in the present study are available upon reasonable request to the authors.

## Acknowledgements

We thank Sebastián Comesaña, Verónica Outeiriño and Inés Pazos of the Centro de Apoio Científico-Técnico á Investigación (CACTI) for their help in sequencing and imaging. We also thank Carlos Solarat, Berta Estévez and Cristina González Abalde for their colony picking skills. We thank Martin R. Wilkins for his contribution to cohort data.

## Funding

This study was funded by grants from the Spanish Ministry of Science and Innovation PI18/01233 to P.E-S, Janssen Pharmaceuticals, and Fundación Contra la Hipertensión Pulmonar. CINBIO has financial support from Xunta de Galicia and the European Union (European Regional Development Fund - ERDF) (PO FEDER ED431G/02). The UK National Cohort of Idiopathic and Heritable PAH is supported by the NIHR Cambridge Biomedical Research Centre, the British Heart Foundation (BHF, SP/18/10/33975), the BHF Cambridge Centre of Cardiovascular Research Excellence, the UK Medical Research Council (MR/K020919/1), the Dinosaur Trust, NIHR Great Ormond Street Hospital Biomedical Research Centre and Great Ormond Street Hospital Charity. M.L-D. was supported by a Xunta de Galicia predoctoral fellowship (ED481A-2018/304). W.L was supported by FS/SBSRF/20/31005. This work was also supported by the Netherlands Cardiovascular Research Initiative: An initiative with the support of the Dutch Heart Foundation (CVON2017-4 DOLPHIN-GENESIS). A.A.R.T. is supported by a BHF Intermediate Clinical Fellowship (FS/18/13/3328).

## Data availability

All data produced in the present study are available upon reasonable request to the authors.

## Notes

### Competing Interest Statement

The authors have declared no competing interest.

### Summary of Updates

In the original submission, we used all the gain-of-function variants in our genotype-phenotype study although some of them were Variants of Uncertain Significance. We have changed this aspect of the study, as we think only those determined pathogenic/likely pathogenic should have been used. Supp. Data and figures 4/5 have been updated. Also, we added all the authors listed in our collaborator subgroup and corrected small errors.

## References

1. Galiè N, Humbert M, Vachiery J-L, Gibbs S, Lang I, Torbicki A, Simonneau G, Peacock A, Noordegraaf AV, Beghetti M, Ghofrani A, Sanchez MAG, Hansmann G, Klepetko W, Lancellotti P, Matucci M, McDonagh T, Pierard LA, Trindade PT, Zompatori M, Hoeper M. 2015 ESC/ERS Guidelines for the diagnosis and treatment of pulmonary hypertension: The Joint Task Force for the Diagnosis and Treatment of Pulmonary Hypertension of the European Society of Cardiology (ESC) and the European Respiratory Society (ERS)Endorsed by: Association for European Paediatric and Congenital Cardiology (AEPC), International Society for Heart and Lung Transplantation (ISHLT). Eur Respir J 2015;46:903–975.

2. Gräf S, Haimel M, Bleda M, Hadinnapola C, Southgate L, Li W, Hodgson J, Liu B, Salmon RM, Southwood M, Machado RD, Martin JM, Treacy CM, Yates K, Daugherty LC, Shamardina O, Whitehorn D, Holden S, Aldred M, Bogaard HJ, Church C, Coghlan G, Condliffe R, Corris PA, Danesino C, Eyries M, Gall H, Ghio S, Ghofrani H-AA, et al. Identification of rare sequence variation underlying heritable pulmonary arterial hypertension. Nat Commun 2018;9:1416.

3. Swietlik EM, Greene D, Zhu N, Megy K, Cogliano M, Rajaram S, Pandya D, Tilly T, Lutz KA, Welch CCL, Pauciulo MW, Southgate L, Martin JM, Treacy CM, Penkett CJ, Stephens JC, Bogaard HJ, Church C, Coghlan G, Coleman AW, Condliffe R, Eichstaedt CA, Eyries M, Gall H, Ghio S, Girerd B, Grünig E, Holden S, Howard L, et al. Bayesian Inference Associates Rare KDR Variants With Specific Phenotypes in Pulmonary Arterial Hypertension. Circ Genomic Precis Med 2020;14:e003155.

4. Zhu N, Swietlik EM, Welch CL, Pauciulo MW, Hagen JJ, Zhou X, Guo Y, Karten J, Pandya D, Tilly T, Lutz KA, Martin JM, Treacy CM, Rosenzweig EB, Krishnan U, Coleman AW, Gonzaga-Juaregui C, Lawrie A, Trembath RC, Wilkins MR, Morrell NW, Shen Y, Gräf S, Nichols WC, Chung WK. Rare variant analysis of 4241 pulmonary arterial hypertension cases from an international consortium implicates FBLN2, PDGFD, and rare de novo variants in PAH. Genome Med 2021;13:.

5. Swietlik EM, Prapa M, Martin JM, Pandya D, Auckland K, Morrell NW, Gräf S. ‘There and Back Again’-Forward Genetics and Reverse Phenotyping in Pulmonary Arterial Hypertension. Genes 2020;11:.

6. Welch CL, Chung WK. Genetics and Genomics of Pediatric Pulmonary Arterial Hypertension. Genes 2020;11:1213.

7. Southgate L, Machado RD, Gräf S, Morrell NW. Molecular genetic framework underlying pulmonary arterial hypertension. Nat Rev Cardiol 2019;doi:10.1038/s41569-019-0242-x.

8. Naiche LA, Harrelson Z, Kelly RG, Papaioannou VE. T-box genes in vertebrate development. Annu Rev Genet 2005;39:219–239.

9. Chapman DL, Garvey N, Hancock S, Alexiou M, Agulnik SI, Gibson-Brown JJ, Cebra-Thomas J, Bollag RJ, Silver LM, Papaioannou VE. Expression of the T-box family genes, Tbx1-Tbx5, during early mouse development. Dev Dyn Off Publ Am Assoc Anat 1996;206:379–390.

10. Gibson-Brown JJ, I Agulnik S null, Silver LM, Papaioannou VE. Expression of T-box genes Tbx2-Tbx5 during chick organogenesis. Mech Dev 1998;74:165–169.

11. Naiche LA, Arora R, Kania A, Lewandoski M, Papaioannou VE. Identity and fate of Tbx4-expressing cells reveal developmental cell fate decisions in the allantois, limb, and external genitalia. Dev Dyn Off Publ Am Assoc Anat 2011;240:2290–2300.

12. Goss AM, Tian Y, Tsukiyama T, Cohen ED, Zhou D, Lu MM, Yamaguchi TP, Morrisey EE. Wnt2/2b and beta-catenin signaling are necessary and sufficient to specify lung progenitors in the foregut. Dev Cell 2009;17:290–298.

13. Arora R, Metzger RJ, Papaioannou VE. Multiple roles and interactions of Tbx4 and Tbx5 in development of the respiratory system. PLoS Genet 2012;8:e1002866.

14. Horie M, Miyashita N, Mikami Y, Noguchi S, Yamauchi Y, Suzukawa M, Fukami T, Ohta K, Asano Y, Sato S, Yamaguchi Y, Ohshima M, Suzuki HI, Saito A, Nagase T. TBX4 is involved in the super-enhancer-driven transcriptional programs underlying features specific to lung fibroblasts. Am J Physiol-Lung Cell Mol Physiol 2018;314:L177–L191.

15. Bongers EMHF, Duijf PHG, van Beersum SEM, Schoots J, van Kampen A, Burckhardt A, Hamel BCJ, Lošan F, Hoefsloot LH, Yntema HG, Knoers Nvam, van Bokhoven H. Mutations in the Human TBX4 Gene Cause Small Patella Syndrome. Am J Hum Genet 2004;74:1239–1248.

16. Ballif BC, Theisen A, Rosenfeld JA, Traylor RN, Gastier-Foster J, Thrush DL, Astbury C, Bartholomew D, McBride KL, Pyatt RE, Shane K, Smith WE, Banks V, Gallentine WB, Brock P, Rudd MK, Adam MP, Keene JA, Phillips JA, Pfotenhauer JP, Gowans GC, Stankiewicz P, Bejjani BA, Shaffer LG. Identification of a recurrent microdeletion at 17q23.1q23.2 flanked by segmental duplications associated with heart defects and limb abnormalities. Am J Hum Genet 2010;86:454–461.

17. Levin ML, Shaffer LG, Lewis RAp6 null, Gresik MV, Lupski JR. Unique de novo interstitial deletion of chromosome 17, del(17) (q23.2q24.3) in a female newborn with multiple congenital anomalies. Am J Med Genet 1995;55:30–32.

18. German K, Deutsch GH, Freed AS, Dipple KM, Chabra S, Bennett JT. Identification of a deletion containing TBX4 in a neonate with acinar dysplasia by rapid exome sequencing. Am J Med Genet A 2019;179:842–845.

19. Karolak JA, Vincent M, Deutsch G, Gambin T, Cogné B, Pichon O, Vetrini F, Mefford HC, Dines JN, Golden-Grant K, Dipple K, Freed AS, Leppig KA, Dishop M, Mowat D, Bennetts B, Gifford AJ, Weber MA, Lee AF, Boerkoel CF, Bartell TM, Ward-Melver C, Besnard T, Petit F, Bache I, Tümer Z, Denis-Musquer M, Joubert M, Martinovic J, et al. Complex Compound Inheritance of Lethal Lung Developmental Disorders Due to Disruption of the TBX-FGF Pathway. Am J Hum Genet 2019;104:213–228.

20. Turro E, Astle WJ, Megy K, Gräf S, Greene D, Shamardina O, Allen HL, Sanchis-Juan A, Frontini M, Thys C, Stephens J, Mapeta R, Burren OS, Downes K, Haimel M, Tuna S, Deevi SVV, Aitman TJ, Bennett DL, Calleja P, Carss K, Caulfield MJ, Chinnery PF, Dixon PH, Gale DP, James R, Koziell A, Laffan MA, Levine AP, et al. Whole-genome sequencing of patients with rare diseases in a national health system. Nature 2020;583:96–102.

21. Hernandez-Gonzalez I, Tenorio J, Palomino-Doza J, Meñaca AM, Ruiz RM, Lago-Docampo M, Gomez MV, Roman JG, Valls ABE, Perez-Olivares C, Valverde D, Carbonell JG, Rodríguez-Monte EGL, del Cerro MJ, Lapunzina P, Escribano-Subias P. Clinical heterogeneity of Pulmonary Arterial Hypertension associated with variants in TBX4. In: West J, editor. PLoS ONE 2020;15:e0232216.

22. Thoré P, Girerd B, Jaïs X, Savale L, Ghigna M-R, Eyries M, Levy M, Ovaert C, Servettaz A, Guillaumot A, Dauphin C, Chabanne C, Boiffard E, Cottin V, Perros F, Simonneau G, Sitbon O, Soubrier F, Bonnet D, Remy-Jardin M, Chaouat A, Humbert M, Montani D. Phenotype and outcome of pulmonary arterial hypertension patients carrying a TBX4 mutation. Eur Respir J 2020;55:1902340.

23. Galambos C, Mullen MP, Shieh JT, Schwerk N, Kielt MJ, Ullmann N, Boldrini R, Stucin-Gantar I, Haass C, Bansal M, Agrawal PB, Johnson J, Peca D, Surace C, Cutrera R, Pauciulo MW, Nichols WC, Griese M, Ivy D, Abman SH, Austin ED, Danhaive O. Phenotype characterisation of TBX4 mutation and deletion carriers with neonatal and paediatric pulmonary hypertension. Eur Respir J 2019;54:.

24. Bragin E, Chatzimichali EA, Wright CF, Hurles ME, Firth HV, Bevan AP, Swaminathan GJ. DECIPHER: database for the interpretation of phenotype-linked plausibly pathogenic sequence and copy-number variation. Nucleic Acids Res 2014;42:D993–D1000.

25. Richards S, Aziz N, Bale S, Bick D, Das S, Gastier-Foster J, Grody WW, Hegde M, Lyon E, Spector E, Voelkerding K, Rehm HL. Standards and guidelines for the interpretation of sequence variants: A joint consensus recommendation of the American College of Medical Genetics and Genomics and the Association for Molecular Pathology. Genet Med 2015;17:405–424.

26. Papaioannou VE. The T-box gene family: emerging roles in development, stem cells and cancer. Dev Camb Engl 2014;141:3819–3833.

27. Stirnimann CU, Ptchelkine D, Grimm C, Müller CW. Structural basis of TBX5-DNA recognition: the T-box domain in its DNA-bound and -unbound form. J Mol Biol 2010;400:71–81.

28. Kulisz A, Simon H-G. An evolutionarily conserved nuclear export signal facilitates cytoplasmic localization of the Tbx5 transcription factor. Mol Cell Biol 2008;28:1553–1564.

29. Murakami M, Nakagawa M, Olson EN, Nakagawa O. A WW domain protein TAZ is a critical coactivator for TBX5, a transcription factor implicated in Holt-Oram syndrome. Proc Natl Acad Sci U S A 2005;102:18034–18039.

30. Zhu N, Gonzaga-Jauregui C, Welch CL, Ma L, Qi H, King AK, Krishnan U, Rosenzweig EB, Ivy DD, Austin ED, Hamid R, Nichols WC, Pauciulo MW, Lutz KA, Sawle A, Reid JG, Overton JD, Baras A, Dewey F, Shen Y, Chung WK. Exome Sequencing in Children With Pulmonary Arterial Hypertension Demonstrates Differences Compared With Adults. Circ Genomic Precis Med 2018;11:e001887.

31. Zweier C, Sticht H, Aydin-Yaylagül I, Campbell CE, Rauch A. Human TBX1 missense mutations cause gain of function resulting in the same phenotype as 22q11.2 deletions. Am J Hum Genet 2007;80:510–517.

32. Postma AV, Van De Meerakker JBA, Mathijssen IB, Barnett P, Christoffels VM, Ilgun A, Lam J, Wilde AAM, Deprez RHL, Moorman AFM. A gain-of-function TBX5 mutation is associated with atypical Holt-Oram syndrome and paroxysmal atrial fibrillation. Circ Res 2008;102:1433–1442.

33. Posch MG, Gramlich M, Sunde M, Schmitt KR, Lee SHY, Richter S, Kersten A, Perrot A, Panek AN, Al Khatib IH, Nemer G, Mégarbané A, Dietz R, Stiller B, Berger F, Harvey RP, Ozcelik C. A gain-of-function TBX20 mutation causes congenital atrial septal defects, patent foramen ovale and cardiac valve defects. J Med Genet 2010;47:230–235.

34. Veitia RA, Birchler JA. Dominance and gene dosage balance in health and disease: why levels matter! J Pathol 2010;220:174–185.

35. Shrivastava S, Kruisselbrink TM, Mohananey A, Thomas BC, Kushwaha SS, Pereira NL. Rare TBX4 Variant Causing Pulmonary Arterial Hypertension With Small Patella Syndrome in an Adult Man. JACC Case Rep 2021;3:1447–1452.

36. Zhu N, Pauciulo MW, Welch CL, Lutz KA, Coleman AW, Gonzaga-Jauregui C, Wang J, Grimes JM, Martin LJ, He H, PAH Biobank Enrolling Centers’ Investigators, Shen Y, Chung WK, Nichols WC. Novel risk genes and mechanisms implicated by exome sequencing of 2572 individuals with pulmonary arterial hypertension. Genome Med 2019;11:69.

37. Whiffin N, Minikel E, Walsh R, O’Donnell-Luria AH, Karczewski K, Ing AY, Barton PJR, Funke B, Cook SA, MacArthur D, Ware JS. Using high-resolution variant frequencies to empower clinical genome interpretation. Genet Med Off J Am Coll Med Genet 2017;19:1151–1158.

38. Karolak JA, Gambin T, Szafranski P, Stankiewicz P. Potential interactions between the TBX4-FGF10 and SHH-FOXF1 signaling during human lung development revealed using ChIP-seq. Respir Res 2021;22:26.

39. Kerstjens-Frederikse WS, Bongers EMHF, Roofthooft MTR, Leter EM, Douwes MJ, Dijk AV, Vonk-Noordegraaf A, Dijk-Bos KK, Hoefsloot LH, Hoendermis ES, Gille JJP, Sikkema-Raddatz B, Hofstra RMW, Berger RMF. TBX4 mutations (small patella syndrome) are associated with childhood-onset pulmonary arterial hypertension. J Med Genet 2013;50:500–506.

