## supplementary data, docx for "First genotype-phenotype study in TBX4 syndrome: gain-of-function mutations causative for lung disease"

[32] www.ipahcohort.com

[33] www.pahbiobank.org

*** Corresponding authors**

Nicholas W. Morrell

University of Cambridge, Department of Medicine

Box 157, Level 5, Addenbrooke's Hospital, Hills Road,

Cambridge, CB2 0QQ, United Kingdom

Tel: (+44) 1223 331666

Stefan Gräf

University of Cambridge, Department of Medicine

Addenbrooke's Hospital, Hills Road,

Cambridge, CB2 0QQ, United Kingdom

Diana Valverde

CINBIO, Universidade de Vigo

Campus Universitario As Lagoas-Marcosende s/n

Vigo 36310, Spain

Tel: (+34) 986 811 953

#### ONLINE DATA SUPPLEMENT

I. **Functional studies- expanded methods**

   A. **eTable 1.** Cloning and sequencing primers.

   B. **eTable 2.** Site-directed mutagenesis primers.

   C. **eTable 3.** qPCR primers.

II. **Variant assessment**

   A. **Functional**

      1. **eFigure 1.** Validation of the overexpression levels of TBX4 at mRNA and protein levels.
      2. **eFigure 2.** *TBX4* variants co-localize to the nuclei independently of their activity.
      3. **eFigure 3.** TBX4 antibody validation for immunofluorescence.
      4. **eFigure 6.** Minigene analysis of *TBX4* splice-site variants.
      5. **eFigure 7.** Results of the luciferase assay for *TBX4* variants annotated as benign.
      6. **eFigure 8.** Optimization of the luciferase assay.
      7. **eFigure 9.** Loss of immunoreactivity of the p.Ile270Ser *TBX4* variant.

   B. ***In silico***

      *1.* **eFigure 4.** Representation of the percentage number of variants with altered classification following functional assessment by luciferase assay.

      *2.* **eFigure 5.** Comparison of the accuracy of different *in silico* prediction tools.

III. **Clinical phenotype**

   A. **Radiological sub-study**

1. **eTable 4.** Reporting proforma for analysis of radiological features.

**B. Histopathology**

**IV. Supplemental References**

### I. Expanded Methods

#### Reporter plasmids cloning

The FGF10 promoter and FGF10 “intronic island” were amplified using Phusion High Fidelity Polymerase (ThermoFisher). The FGF10 promoter was digested with XhoI + NheI and the FGF10 “intronic island” with SalI + NheI (NZYtech). Then, both were ligated for 1 hour at 22 °C following a 5:1 insert:vector ratio with pGL3 (Promega) and pmirGLO (Promega) respectively using a T4 ligase (Canvax).

The minipGL3 and minipmirGLO TBOX promoters were generated by annealing overlapping primers with x3 TBOX regions flanked by restriction enzyme sites. We annealed them by diluting 100 ng of each in 20 µL, increasing the temperature up to 95 °C for 3 minutes, and letting it cool on the bench to room temperature. Then, the products were digested with NheI + EcoRI/SalI (NZYtech) and ligated into pGL3 and pmirGLO.

For the bacterial transformation, we used 5 µl of the ligation product and 95 µl of NZYStar competent cells (NZYtech) following the manufacturer’s protocol. Transformants were screened by colony PCR using NZYTaq Green Mastermix (NZYtech) with the appropriate combination of sequencing primers from the eTable 1. We diluted the colonies in 5 µL of PBS and used 1 µL for the PCR in a total volume of 25 µL. PCR products were fractionated in a 2 % agarose gel and bands were visualized in a GelDoc EQ (BioRad). Finally, PCR bands were excised and purified prior to submission for  Sanger sequencing in the *Centro de Apoio Científico Técnico á Investigación* (CACTI) of the University of Vigo.

**Site-directed mutagenesis**

We carried out site-directed mutagenesis for each of the variants using the primers listed in eTable 2 in the TBX4-MYC-DKK (Origene #RC217451) and the minigene constructs. The reaction was performed using NZYMut site-directed mutagenesis kit (NZYtech) following the manufacturer's protocol with slight modifications: elongation temperature was increased to 67 °C and all the reactions were supplemented with 5 % DMSO. To generate indels, we used Q5 site-directed mutagenesis kit (New England Biolabs) following the manufacturer's protocol.

The mutant screening was undertaken by colony PCR as described in the previous section. Positive colonies were expanded in 3 mL of LB media supplemented with the appropriate antibiotic and then harvested to extract plasmid DNA using NZYminiprep (NZYtech). Plasmid DNA was quantified using a Nanodrop 3000 (ThermoFisher). PCR products were Sanger sequenced to confirm the *TBX4* mutations.

**Cell culture**

We cultured HeLa cells (ATCC) in DMEM (Corning) supplemented with 1 % streptomycin/penicillin and 10 % Fetal Bovine Serum (FBS).

**Luciferase assay**

For the luciferase assay, HeLa cells were grown in 24 well plates until achieving 80-90 % confluence. Transfection was carried out using Lipofectamine 3000 (ThermoFisher) following the manufacturer's protocol, after 24 hours we removed the transfection media and added full growth media. 24 hours later cells were washed and harvested for analysis using the DualGlo® Luciferase Assay System (Promega) according to the manufacturer's instructions. The luciferase results were read in an EnVision 2100 multilabel reader (Perkin Elmer) using white half-area 96-well plates (Greiner). Firefly luciferase results were normalized against *Renilla*, and the data were represented as a fold change of the WT.

**Optimization of the luciferase assay**

To optimize the reporter to use and the quantity of TBX4-overexpression plasmids, we transfected cells in 96-well plates with 40 ng of a reporter (empty pGL3/pmirGLO, miniTBX-pGL3 or miniTBX-pmirGLO) and either 40/30/20/10/0 ng of the WT TBX4 overexpression plasmid or 40 ng of a predicted Pathogenic variant (p.Val103_Lys104insGlu) and a nonsense variant control (p.Tyr127Ter). All the conditions were scaled proportionally to conduct the experiments in 24-well plates.

**Real-Time quantitative PCR**

We cultured HeLa cells in 24-well plates until confluence, then transfected them with 100 ng of each of the variants (WT, p.Pro98Arg, p.Met144Ile, p.Arg250Trp, p.Arg261Gln, and p.Ile270Ser) using lipofectamine 3000 following the manufacturer's protocol. Fresh medium was added the next day and the cells were harvested 24 hours later. RNA extraction was carried out using the NZY Total RNA Isolation Kit (NZYtech) following the manufacturer's protocol. We used 100 ng of RNA for retrotranscription using NZY M-MuLV First-Strand cDNA synthesis kit (NZYtech). Real-Time quantitative PCR (qPCR) was carried out using PowerUp SYBR Green Master Mix (ThermoFisher), 1 µL of 1:10 cDNA dilution, and the primers shown in eTable 3. The reaction was performed using a total volume of 15 µL in a Step-One Plus Real-Time PCR system (ThermoFisher), cycling conditions were as follows: 50 °C for 2 minutes, 95 °C for 2 minutes, 40 cycles of 95 °C for 15 seconds and 30 seconds at 60 °C; followed by a melting curve.

To normalize the expression, we used the -ΔCT method using *YWHAZ* and *ALAS1* as reference genes.

**Western blot**

Hela cells were grown in 6-well plates until 80-90% confluence. Then, we transfected the cells with 500 ng of plasmid DNA for each of the chosen variants (WT, p.Pro98Arg, p.Met144Ile, p.Arg250Trp, p.Arg261Gln, and p.Ile270Ser) using lipofectamine 3000 (ThermoFisher) following the manufacturer's protocol. After 24 hours, we changed the media, and the following day we extracted protein for each of the variants using RIPA buffer supplemented with protease inhibitors (Sigma).

We prepared 20 µg of protein in Laemlii's sample buffer (BioRad) containing 5 % Β-mercaptoethanol (Sigma) and heated it at 95 °C for 5 minutes. Proteins were separated by SDS-Page using a 12 % mini-Protean TGX precast gel (BioRad). After electrophoresis, we transferred the proteins to a PVDF membrane using a Trans-Blot Turbo Transfer pack (BioRad) in a Trans-Blot Turbo system (BioRad) for 7 minutes at 1.3 A. We then incubated the membrane for 1 hour in a blocking buffer composed of 5 % w/v nonfat milk in Tris-buffered saline (TBS) containing 0.2 % Tween20 (TBS-T). Immunoblotting was carried out by incubating the membrane at room temperature with a 1:1000 dilution of anti-Flag-HRP antibody (Abcam #ab49763) for 1 hour. We then washed the membrane with TBS and developed the blot using Clarity Western ECL substrate (BioRad).

We treated the membrane with Restore Plus Stripping Buffer (ThermoFisher) for 5 minutes, washed it in TBS, repeated the blocking step, and incubated it at 4 °C overnight with a 1:500 dilution of anti-TBX4 antibody (Sigma #AV33739). Then, we washed the membrane three times in blocking buffer and incubated with a 1/10000 dilution of Goat anti-Rabbit IgG (Abcam #ab205718) for 1 hour at room temperature before developing the blot in the same way as before. Imaging was carried out in a ChemiDoc™ (Bio-Rad) digital camera-based imaging system. We used total protein as loading control by staining the membrane with Coomassie Brilliant Blue.

**Immunofluorescence**

We cultured HeLa cells in 6 well plates with a 24x24 mm glass coverslip until 80-90 % confluence was achieved. Transfection was carried out with the same conditions and for the same variants as stated in the western blot protocol. Cells were washed 3 times in PBS before being fixed with 4 % formalin for 10 minutes at 37 ºC. After washing the cells six times in PBS, we proceeded to permeabilize them in PBS+BSA 1 % (w/v) containing 0.1 % Triton X-100 (v/v). Then, we blocked them in PBS + BSA 2 % (blocking buffer) for 1 hour at room temperature. We incubated the cells with the primary antibodies overnight in blocking buffer, washed three times in blocking buffer for 5 minutes, and incubated with the secondary antibodies and 4', 6-diamidino-2-phenylindole dihydrochloride (DAPI) (1 µg/mL) in blocking buffer for 1 hour in the dark. Finally, we washed the coverslips three times for 5 minutes in PBS and mounted them in ProLong Diamond Antifade Mountant (ThermoFisher). Images were acquired using a Leica DMI6000 inverted microscope with an integrated confocal module SP5 (Leica Microsystems). All the images were processed with ImageJ (v.1.8.0). For co-localization analysis, we used the  EzColocalization plugin(1).

The following antibodies and dilutions were used: anti-TBX4 (Sigma #AV33739, 1:500), anti-pancadherin (abcam #ab22744, 1:500), Alexa Fluor 488-conjugated goat anti-rabbit (ThermoFisher #A-11008, 1:1000) and Alexa Fluor 594-conjugated goat anti-mouse (ThermoFisher #A-11005, 1:1000). To validate the TBX4 antibody, we compared the results with TBX4-GFP (Origene #RG217451) expression.

**Minigene assay**

We used Phusion High Fidelity Polymerase (ThermoFisher) to amplify 2 regions of the *TBX4* gene, one including exon 5 (TBX4_e5_pSPL3), and another including exons 6 and 7 (TBX4_e6_7_pSPL3) using the primers described in eTable 1. We purified the amplicons, digested them with EcoRI/XhoI + NheI (NZYtech), and cloned the products into the exon trapping p.SPL3 vector (ThermoFisher). Transformants were selected with ampicillin and confirmed by Sanger sequencing using colony PCR.

Mutants were generated by site-directed mutagenesis with the primers described in eTable 2, 2.5 µg of each of the variants were used for the transfection of HeLa cells with Lipofectamine 2000 (ThermoFisher) following the manufacturer's recommendations. After 48 hours, we extracted RNA and carried out an RT-PCR in the same way as stated in the qPCR protocol. We then used 2 µL of cDNA for a PCR using Phusion High Fidelity Polymerase (ThermoFisher), the primers SA2 (5'-ATCTCAGTGGTATTTGTGAGC-3') and SD6 (5'-TCTGAGTCACCTGGACAACC-3) with the following thermocycling conditions: 98 ºC for 3 minutes, 35 cycles of 10 seconds at 98 ºC, 30 seconds at 58 ºC and 30 seconds at 72 ºC, followed by a final extension at 72 ºC for 7 minutes. Finally, we separated the PCR products by gel electrophoresis and imaged them. PCR products were submitted for Sanger sequencing to confirm the presence/absence of the exons.

**Variant classification**

We assessed variant pathogenicity according to the American College of Medical Genetics and Genomics (ACMG) guidelines (2) using the VarSome Clinical tool followed by manual curation. The BP1 criterion (missense variant in a gene for which primarily truncating variants are known to cause disease) was removed whenever automatically applied by VarSome; as illustrated by this study, both truncating and missense variants are causative for TBX4-associated disease phenotypes. BS2 (variant for highly penetrant condition seen in healthy individuals) was also removed as the penetrance of *TBX4* sequence variants is suspected to be low, albeit no accurate estimates exist to date. Instead, we used the BS1 criterion (allele frequency is greater than expected for the disorder) whenever applicable. We calculated the maximum tolerated allele frequency plausible for a *TBX4* pathogenic variant using the statistical framework developed by Whiffin et al. (3) available from DECIPHER. This was estimated at $5.00 \times 10^{-8}$ using a dominant inheritance model with idiopathic pulmonary arterial hypertension (IPAH) prevalence of 1 in 1.000.000 (Orphanet), TBX4 lung disease penetrance of 0.5, and maximum allelic contribution of *TBX4* to the PAH genetic architecture of approximately 5% taking into account both pediatric- and adult-onset cohorts (4–7). Where the ethnicity of individual cases was known, population frequency data (gnomAD) were filtered accordingly. The PS3/BS3 criterion (+/- damaging effect on protein function or splicing) was applied to reclassify functionally assessed variants (supplementary data, xlsx).

**Statistical analysis and data visualization**

For the luciferase results, we first performed a Shapiro-Wilk test to assess normality, followed by one-way ANOVA with a Bonferroni correction for multiple comparisons using the ggpubr package in R. Box plots and dot plots were generated using the R package ggplot2 (8).

**eTable 1. Cloning and sequencing primers.**

| Primer ID | Sequence 5' - 3' | Annealing T (°C) |
|---|---|---|
| FGF10promF_XhoI | AAACTCGAGGCACCAACATCCATAACTCC | 66 |
| FGF10promR_NheI | AAAGCTAGCCCAATATGGAGGTCAAACGC | 66 |
| FGF10_INT_NheI_R | AAAGCTAGCCGTGTAGAGGTGCTTTTAAAGTG | 65 |
| FGF10_INT_SalI_F | AAAGTCGACGCAAACCTCAAAAGAACAGC | 65 |
| miniTBX4_pmirGLO_F | CTAGCAGGTGTGAAGGTGTGAAGGTGTGAG | 55 |
| miniTBX4_pmirGLO_R | TCGACTCACACCTTCACACCTTCACACCTG | 55 |
| miniTBX4_pGL3_F | CTAGCAGGTGTGAAGGTGTGAAGGTGTGAG | 55 |
| miniTBX4_pGL3_R | AATTCTCACACCTTCACACCTTCACACCTG | 55 |
| TBX4_exon6-7_F | AAAGGATCCTCCCCAAGGAGGGTAGTGAG | 58 |
| TBX4_exon6-7_R | AAAGCTAGCGCCCAGCTGTGATCCCTAAC | 58 |
| TBX4_exon5_F | ACAGAATTCGCACCCTGGACTTTTGCTGA | 58 |
| TBX4_exon5_R | AAAGCTAGCCCTGTCTCTGAAGGCACCAG | 58 |
| pgl3_lucN_R | CCTTATGCAGTTGCTCTCC | 55 |
| EBV-rev | GTGGTTTGTCCAAACTCATC | 55 |
| T7_F | TAATACGACTCACTATAGGG | 55 |
| TBX4_3F | TGGAAGAAGTTCCACGAGGC | 55 |
| TBX4_3F_anti | GCCTCGTGGAACTTCTTCCA | 55 |
| TBX4_4F | AGTGATGACAGTGACCTGCG | 55 |
| TBX4_4R | GGGCTTCCAGATAGGATCGC | 55 |
| TBX4_2F | CAGCTATAGCGTGCAGACGA | 55 |
| TBX4_2F_anti | TCGTCTGCACGCTATAGCTG | 55 |
| TBX4_2R | CCGTCAGTCCAGTTCTCCAC | 55 |

**eTable 2. Site-directed mutagenesis primers.**

| Primer ID | Sequence 5' - 3' | Annealing T (°C) |
|---|---|---|
| Glu9Lys_anti | GCCTCCTCGCTCTTGGACAGGCCCTTATCC | 67 |
| Glu9Lys_sense | GGATAAGGGCCTGTCCAAGAGCGAGGAGGC | 67 |
| Ala35Val_anti | GCTGAGGCCCGGCGCTACCAGCGCGGGCTCGG | 67 |
| Ala35Val_sense | CCGAGCCCGCGCTGGTAGCGCCGGGCCTCAGC | 67 |
| Ile64Phe_anti | CCTTGATGTTCTCGAAGGTCTGCTCCGCGG | 67 |
| Ile64Phe_sense | CCGCGGAGCAGACCTTCGAGAACATCAAGG | 67 |
| Trp77Arg_anti | TCGTGGAACTTCTTCCGGAGCTCCTTCTCATGC | 67 |
| Trp77Arg_sense | GCATGAGAAGGAGCTCCGGAAGAAGTTCCACGA | 67 |
| Glu86Gln_anti | AGTGATGATCATCTGGGTGCCCGCCTCG | 67 |
| Glu86Gln_sense | CGAGGCGGGCACCCAGATGATCATCACT | 67 |
| Glu86Lys_anti | CCTTAGTGATGATCATCTTGGTGCCCGCCTCGTG | 67 |
| Glu86Lys_sense | CACGAGGCGGGCACCAAGATGATCATCACTAAGG | 67 |
| Met96Lys_anti | GTAGCTGGGGAACTTCCTCCTGCCAGC | 67 |
| Met96Lys_sense | GCTGGCAGGAGGAAGTTCCCCAGCTAC | 67 |
| Pro98Ala_anti | TTACCTTGTAGCTGGCGAACATCCTCCTGCC | 67 |
| Pro98Ala_sense | GGCAGGAGGATGTTCGCCAGCTACAAGGTAA | 67 |
| Pro98Leu_anti | CTTTTACCTTGTAGCTGAGGAACATCCTCCTGCCA | 67 |
| Pro98Leu_sense | TGGCAGGAGGATGTTCCTCAGCTACAAGGTAAAA G | 67 |
| Pro98Arg_anti | CTTTTACCTTGTAGCTGCGGAACATCCTCCTGCC | 67 |
| Pro98Arg_sense | GGCAGGAGGATGTTCCGCAGCTACAAGGTAAAAG | 67 |
| Tyr100Cys_anti | CTGTGACTTTTACCTTGCAGCTGGGGAACATCCTC | 67 |
| Tyr100Cys_sense | GAGGATGTTCCCCAGCTGCAAGGTAAAAGTCACA G | 67 |
| Lys103_Val104ins_sen se | CCCCAGCTACAAGGTAAAAGAAGTCACAGGCATG AACCCC | 67 |

| | | |
|---|---|---|
| Lys103_Val104ins_anti | GGGGTTCATGCCTGTGACTTCTTTTACCTTGTAGC TGGGG | 67 |
| Gly106Ser_anti | TGGGGTTCATGCTTGTGACTTTTACCTTGTAGCTG G | 67 |
| Gly106Ser_sense | CCAGCTACAAGGTAAAAGTCACAAGCATGAACCC CA | 67 |
| Tyr113Cys_anti | TGTCAATCAGCAGGATACACTTGGTCTTGGGGTTC | 67 |
| Tyr113Cys_sense | GAACCCCAAGACCAAGTGTATCCTGCTGATTGAC A | 67 |
| Tyr127Ser_anti | GTTGTCACAGAACTTGGAGCGATGGTCATCGGC | 67 |
| Tyr127Ser_sense | GCCGATGACCATCGCTCCAAGTTCTGTGACAAC | 67 |
| Tyr127Asn_sense | CTGCCGATGACCATCGCAACAAGTTCTGTGACAA C | 67 |
| Tyr127Asn_sense | GTTGTCACAGAACTTGTTGCGATGGTCATCGGCA G | 67 |
| Tyr127Ter_sense | GCCGATGACCATCGCTAGAAGTTCTGTGACAACA AATGG | 67 |
| Tyr127Ter_anti | CCATTTGTTGTCACAGAACTTCTAGCGATGGTCAT CGGC | 67 |
| Met144Ile_anti | CAGCCTTCCTGGAATGGCTGGCTCAGC | 67 |
| Met144Ile_sense | GCTGAGCCAGCCATTCCAGGAAGGCTG | 67 |
| Pro152Leu_anti | GTGGCAGGAGAATCCAGGTGGACATACAGCC | 67 |
| Pro152Leu_sense | GGCTGTATGTCCACCTGGATTCTCCTGCCAC | 67 |
| His177Tyr_anti | CCAAAGGGGTCCAGGTAGTTGTTTGTCAGCTTCA GC | 67 |
| His177Tyr_sense | GCTGAAGCTGACAAACAACTACCTGGACCCCTTT GG | 67 |
| Leu186Arg_anti | GGTACTTGTGCATAGAGTTGCGGATGATATGGCC AAAGG | 67 |
| Leu186Arg_sense | CCTTTGGCCATATCATCCGCAACTCTATGCACAAG TACC | 67 |
| His190Pro_anti | CGCGGCTGGTACTTGGGCATAGAGTTGAGGA | 67 |
| His190Pro_sense | TCCTCAACTCTATGCCCAAGTACCAGCCGCG | 67 |
| Val218Met_anti | AGGTCTCTGGGAACATGTGGGTGCAGAAAGC | 67 |

| | | |
|---|---|---|
| Val218Met_sense | GCTTTCTGCACCCACATGTTCCCAGAGACCT | 67 |
| Ser226Tyr_anti | GGTAGGAGGTCACATAGATGAAGGAGGTCTCTGG G | 67 |
| Ser226Tyr_sense | CCCAGAGACCTCCTTCATCTATGTGACCTCCTACC | 67 |
| Ile235Ser_anti | CTCAATTTTCAGCTGGGTGCTCTTGTGATTCTGGT AGG | 67 |
| Ile235Ser_sense | CCTACCAGAATCACAAGAGCACCCAGCTGAAAATT GAG | 67 |
| Gly248Val_anti | CACTGCCCCGGAATACCTTGGCAAAAGGG | 67 |
| Gly248Val_sense | CCCTTTTGCCAAGGTATTCCGGGGCAGTG | 67 |
| Arg250Trp_anti | CTGTCATCACTGCCCCAGAATCCCTTGGCAAAAG | 67 |
| Arg250Trp_sense | CTTTTGCCAAGGGATTCTGGGGCAGTGATGACAG | 67 |
| Arg250Gln_anti | CTGTCATCACTGCCCTGGAATCCCTTGGCAA | 67 |
| Arg250Gln_sense | TTGCCAAGGGATTCCAGGGCAGTGATGACAG | 67 |
| Arg261Gln_anti | CTTTGCTCTGCAGTTGGGCCACACGCAGG | 67 |
| Arg261Gln_sense | CCTGCGTGTGGCCCAACTGCAGAGCAAAG | 67 |
| Ile270Ser_anti | CTCATGATGCTTTTGGAACTCACGGGGTATTCTTT GC | 67 |
| Gly295Ala_anti | CCTGGTGGGTGGCGAGCAGGGGG | 67 |
| Gly295Ala_sense | CCCCCTGCTCGCCACCCACCAGG | 67 |
| Ile270Ser_sense | GCAAAGAATACCCCGTGAGTTCCAAAAGCATCAT GAG | 67 |
| Asp329Tyr_anti | TAGAAGAGGCTTGAGTACCTCTGGGTGGGAAAG | 67 |
| Asp329Tyr_sense | CTTTCCCACCCAGAGGTACTCAAGCCTCTTCTA | 67 |
| Asp341His_anti | CGGGTACCGTGTCGTCTTTTCAGGCAGTG | 67 |
| Asp341His_sense | CACTGCCTGAAAAGACGACACGGTACCCG | 67 |
| Arg352Leu_anti | GGGCTTCCAGATAGGATAGCTTGCAAGGTAAGTC C | 67 |
| Arg352Leu_sense | GGACTTACCTTGCAAGCTATCCTATCTGGAAGCCC | 67 |
| Ala357Val_anti | CCCACCGAAGAGGGGACTTCCAGATAGGATC | 67 |

| | | |
|---|---|---|
| Ala357Val_sense | GATCCTATCTGGAAGTCCCCTCTTCGGTGGG | 67 |
| Arg368Cys_anti | GAGGGGGGGAACAGAAATAGTGATCCTCCC | 67 |
| Arg368Cys_sense | GGGAGGATCACTATTTCTGTTCCCCCCCTC | 67 |
| Tyr382Ser_anti | CACCTCACTGCAGGAGGAGGGGCTCAG | 67 |
| Tyr382Ser_sense | CTGAGCCCCTCCTCCTGCAGTGAGGTG | 67 |
| Ser395Pro_anti | CGGGCCCTGAACCTGGGTACATACATGCTTCTC | 67 |
| Ser395Pro_sense | GAGAAGCATGTATGTACCCAGGTTCAGGGCCCG | 67 |
| Glu400Lys_anti | CCCCGGCAATCTTGGGCCCTGAACC | 67 |
| Glu400Lys_sense | GGTTCAGGGCCCAAGATTGCCGGGG | 67 |
| Gly403Arg_anti | CCCCAGACACCCTGGCAATCTCGGG | 67 |
| Gly403Arg_sense | CCCGAGATTGCCAGGGTGTCTGGGG | 67 |
| Pro425Gln_anti | GCTATAGCTGGTGTACTGCGACACTGAAGTCC | 67 |
| Pro425Gln_sense | GGACTTCAGTGTCGCAGTACACCAGCTATAGC | 67 |
| Met451Val_anti | CCGCGGCATCACGGTGGTGGCGG | 67 |
| Met451Val_sense | CCGCCACCACCGTGATGCCGCGG | 67 |
| Asn475His_anti | GAGACTGGGAGAGCTGATCGTAGACACTAAAGTG GG | 67 |
| Asn475His_sense | CCCACTTTAGTGTCTACGATCAGCTCTCCCAGTCT C | 67 |
| Glu515Lys_anti | GGTTTGAGAGTAGAGAAACTTATTGGCAGCATTTA GATGTGGC | 67 |
| Glu515Lys_sense | GCCACATCTAAATGCTGCCAATAAGTTTCTCTACT CTCAAACC | 67 |
| Gln531Arg_anti | CCATTCCTGAATGGTACGGTAAGGAAGATTCTCG G | 67 |
| Gln531Arg_sense | CCGAGAATCTTCCTTACCGTACCATTCAGGAATGG | 67 |
| 702mut_anti | GCAGTGGGGCAGTGGCTGTATCTTGTGATTCTGG TAGG | 67 |
| 702mut_sense | CCTACCAGAATCACAAGATACAGCCACTGCCCCA CTGC | 67 |
| 792-1_anti | CGGGGTATTCTTTGGTGAAGGGGTGGGGA | 67 |

| | | |
|---|---|---|
| 792-1_sense | TCCCCACCCCTTCACCAAAGAATACCCCG | 67 |
| 1021+1GA_anti | GACCAGGAGAGCCCTATCTCGTCTTTTCAGGC | 67 |
| 1021+1GA_sense | GCCTGAAAAGACGAGATAGGGCTCTCCTGGTC | 67 |
| Q5_Ser167del_F | CTTCCAGAAGCTGAAGCTGACAAAC | 69 |
| Q5_Ser167del_R | ACCAGCTGCCGCATCCAG | 69 |
| Q5_Lys172_Leu173del_F | GAAGCTGACAAACAACCACC | 65 |
| Q5_Lys172_Leu173del_R | TGGAAGGAGACCAGCTGC | 65 |
| Q5_Lys191_Tyr192del_F | CAGCCGCGGCTCCACATC | 69 |
| Q5_Lys191_Tyr192del_R | GTGCATAGAGTTGAGGATGATATGGC | 69 |
| Q5_Phe224del_F | ATCTCTGTGACCTCCTAC | 60 |
| Q5_Phe224del_R | GGAGGTCTCTGGGAACAC | 60 |
| Q5_Phe224_Ser229dup_F | cagagatgaAGGAGGTCACAGAGATGAAG | 59 |
| Q5_Phe224_Ser229dup_R | cagagatgaAGGAGGTCACAGAGATGAAG | 59 |

**eTable 3. qPCR primers.**

| Primer ID | Sequence 5' - 3' | Annealing T (°C) |
|---|---|---|
| TBX4_qPCR_F | CTTTCCCACCCAGAGGGACT | 60 |
| TBX4_qPCR_R | GGGGCTTCCAGATAGGATCG | 60 |
| ALAS_qPCR_F | AGTGTGAAAACCGATGGAGG | 60 |
| ALAS_qPCR_R | CGATCATACTGAAAAGTGGAAACAG | 60 |
| YWHAZ_qPCR_F | ATGCAACCAACACATCCTATC | 60 |
| YWHAZ_qPCR_R | GCATTATTAGCGTGCTGTCTT | 60 |

### II. Variant assessment

#### A. Functional

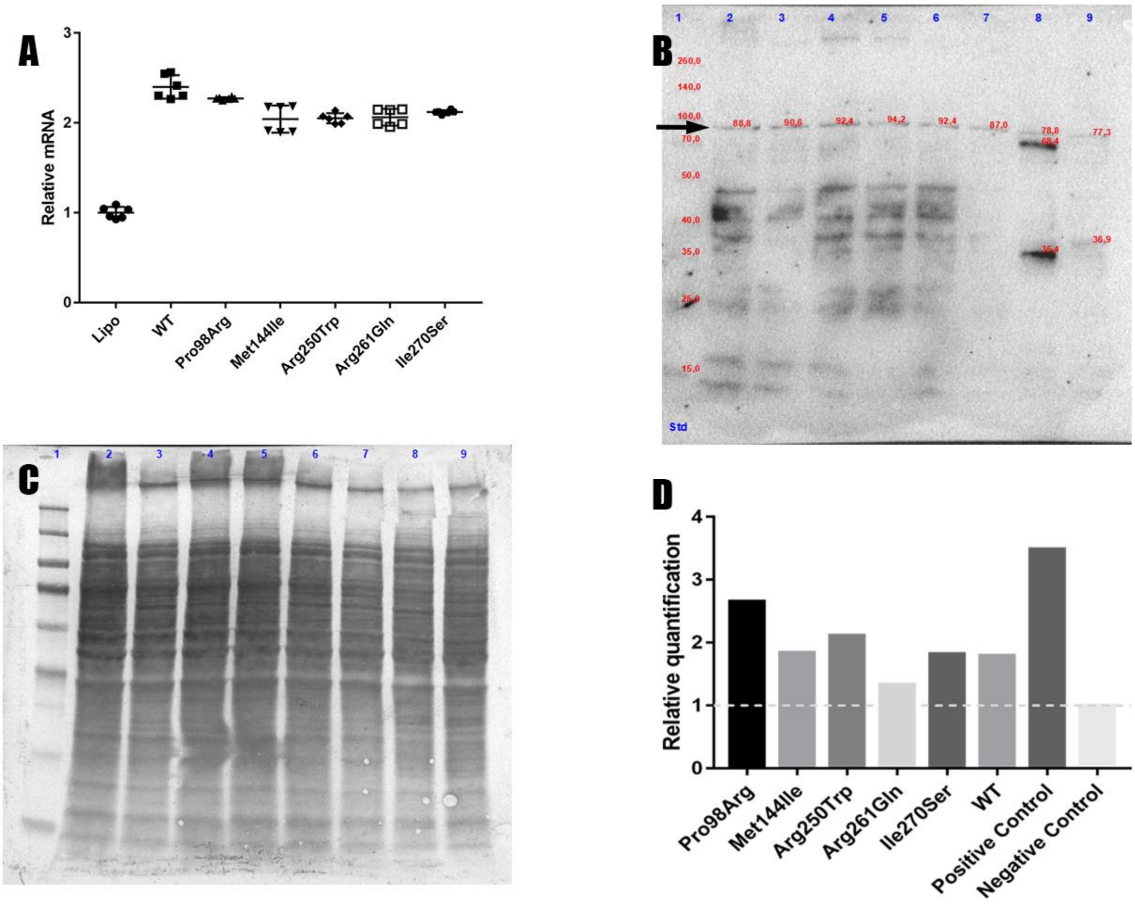

**eFigure 1. Validation of the overexpression levels of TBX4 at mRNA and protein levels.** A) qPCR results after transfection with the TBX4 constructs. *TBX4* mRNA expression of the wild-type and variants are 2-fold higher than the lipofectamine control after normalization. Data are mean ± standard deviation. B) FlagHRP western blot of HeLa cells after transfection under the same conditions as for the qPCR. All the variants show a band of 80-90kDa corresponding to TBX4 after double sumoylation. A 70 kDa band in the positive control may be TBX4 after with single sumoylation. C) Coomassie staining of the PVDF membrane used in the western blot. All the samples were loaded in similar levels. D) Quantification of the western blot using Coomassie staining as a loading control, each bar represents the quantification for the lane above it. The variant p.Pro98Arg (LoF) showed the highest level and p.Arg261Gln (GoF) the lowest with just 1.3 fold. Data are presented as band intensity/lane protein intensity.

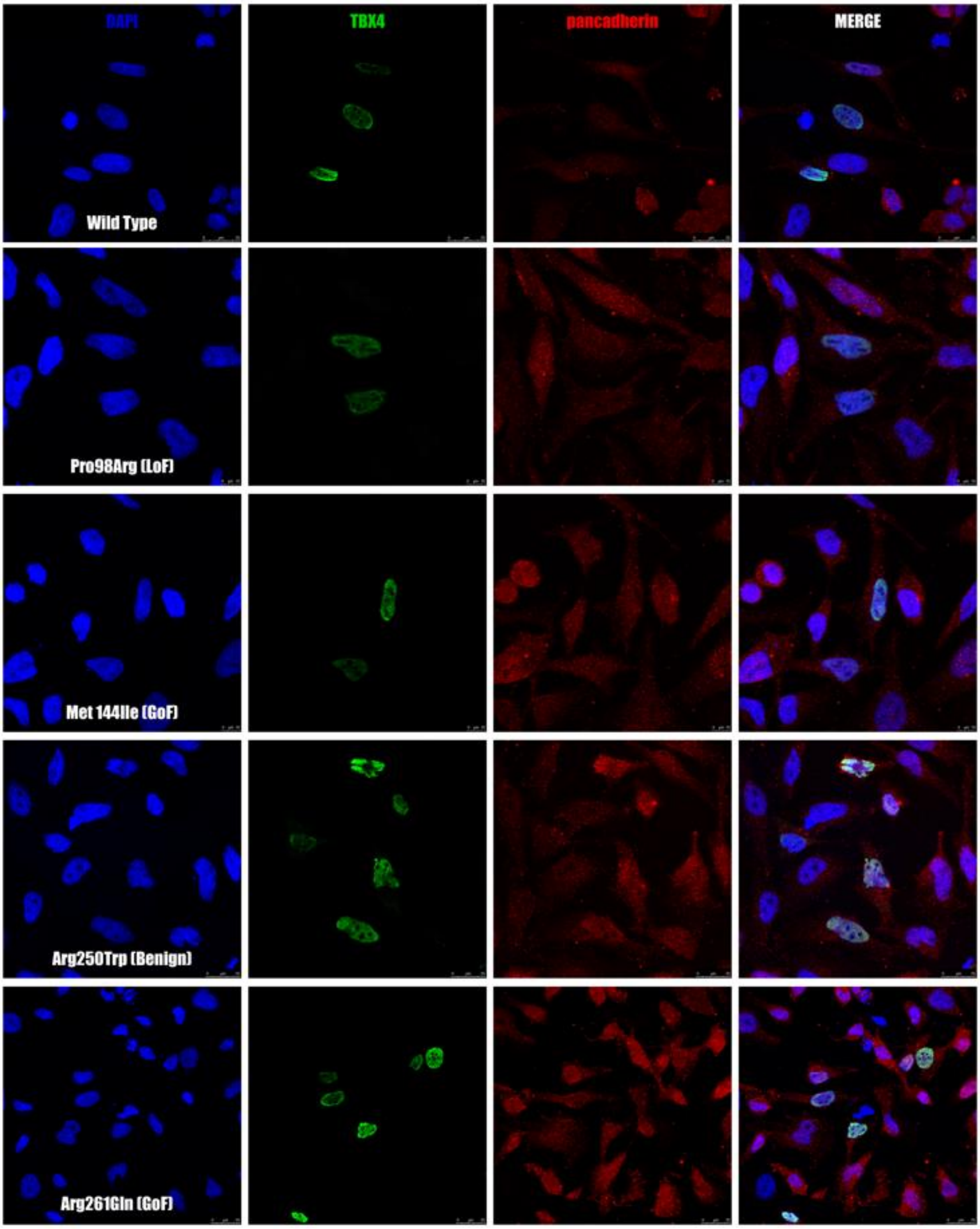


**eFigure 2. *TBX4* variants co-localize to the nuclei independently of their activity.** A) Immunofluorescence for the different *TBX4* variants marked with: nuclei (blue/DAPI), anti-TBX4 (green/alexa488) and anti-pan-cadherin (red/alexa594). All the variants colocalize with DAPI (n = 2).

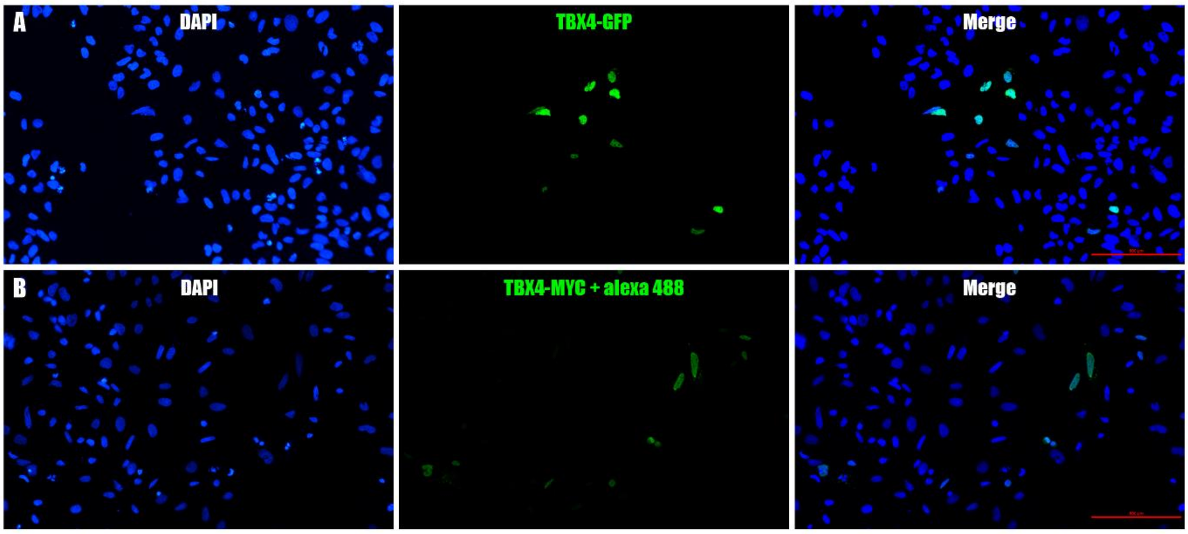


**eFigure 3. TBX4 antibody validation for immunofluorescence.** Cells were transfected with TBX4-GFP (A) and TBX4-MYC-HA (B). We imaged the cells in a fluorescence microscope to confirm that we had the same pattern between GFP and the antibody.

#### Minigene assay

Using hybrid minigenes encoding exon 5, we confirmed that c.702+1G>A induces the skipping of exon 5. The absence of this exon does not induce a frameshift, so the resulting transcript could yield a protein of 494 amino acids lacking 51 amino acids in the T-BOX domain, which taking into account our indel data should make the protein non-functional. The minigenes including exons 6 and 7 showed that the variant c.792-1G>C does not affect the correct processing of these *TBX4* exons, while the recurrent variant c.1021+1G>A induces a double exon skipping event where both exon 6 and 7 are skipped in the mutated constructs. No frameshift is induced in this double skipping, but the resulting protein would have a reduced length of 271 amino acids if not degraded.

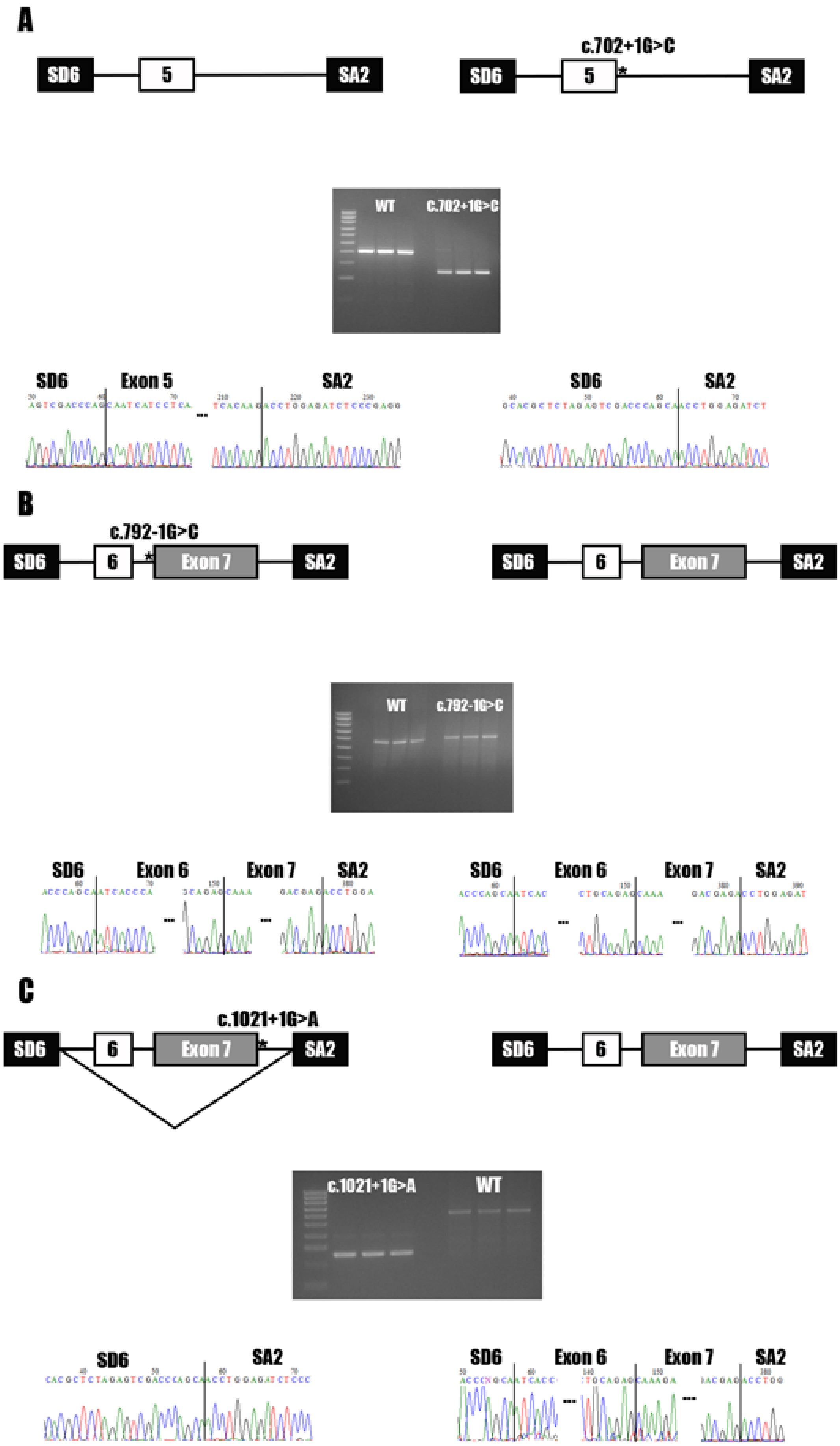
A
SD6
5
SA2
c.702+1G>C
SD6
5
SA2
WT
C.702+1G>C
SD6
Exon 5
SA2
SD6
SA2
B
c.792-1G>C
SD6
6
Exon 7
SA2
SD6
6
Exon 7
SA2
WT
c.792-1G>C
SD6
Exon 6
Exon 7
SA2
SD6
Exon 6
Exon 7
SA2
C
c.1021+1G>A
SD6
6
Exon 7
SA2
SD6
6
Exon 7
SA2
c.1021+1G>A
WT
SD6
SA2
SD6
Exon 6
Exon 7
SA2

**eFigure 6. Minigene assay of *TBX4* splice-site variants.** A) c.702+1G>C induces the skipping of exon 5. B) c.792-1G>C appears benign as splicing was identical to the wild-type. C) c.1021+1G>A induces a double exon skipping, as the minigene lost the exons 6 and 7 that were encoded.

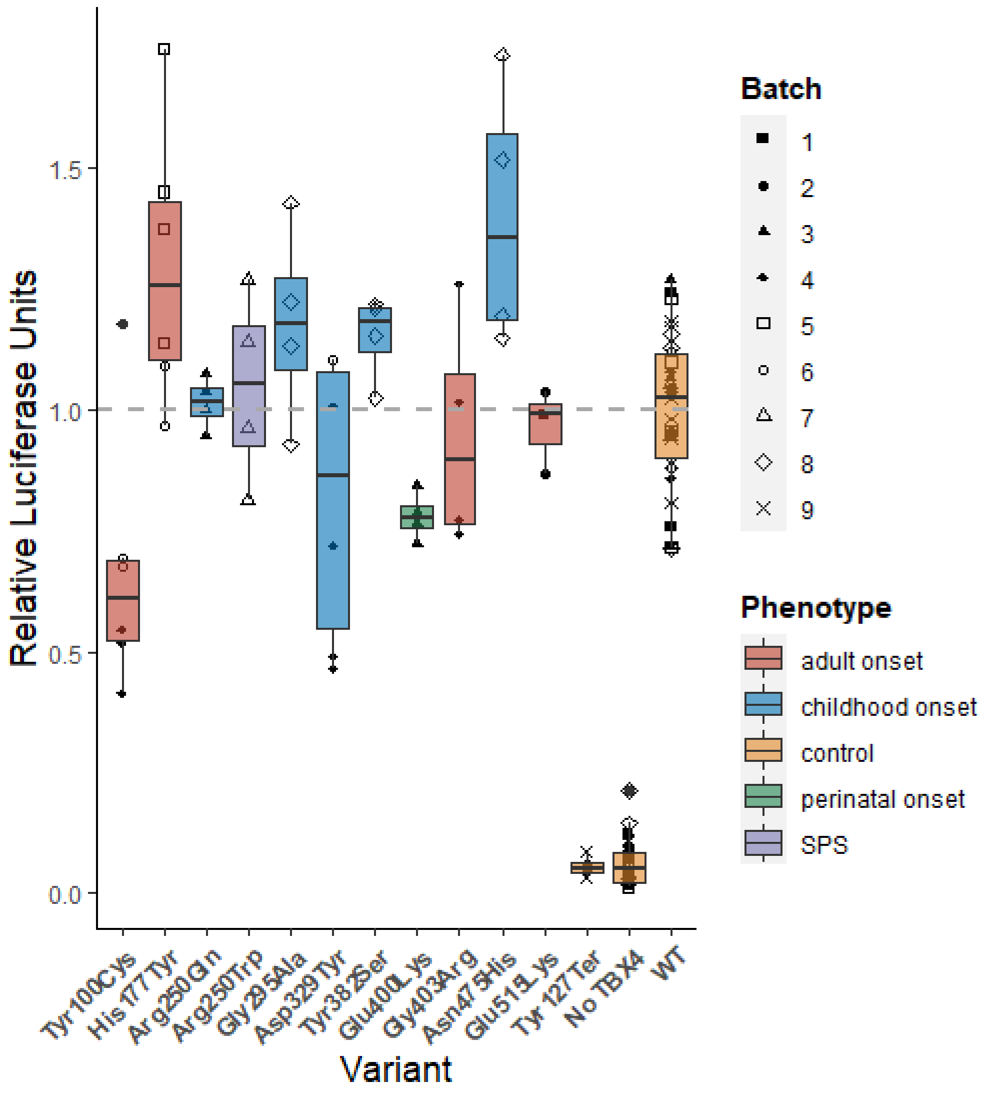


**eFigure 7. Results of the luciferase assay for *TBX4* variants annotated as benign.** We calculated the Firefly:Renilla ratio for each sample and normalized the data using the WT treatment, which was designated as 1. Data are shown as median ± interquartile ranges for at least 4 different replicate experiments. None of these

variants were statistically significant when compared with WT after One-Way ANOVA with a Bonferroni post-hoc correction.

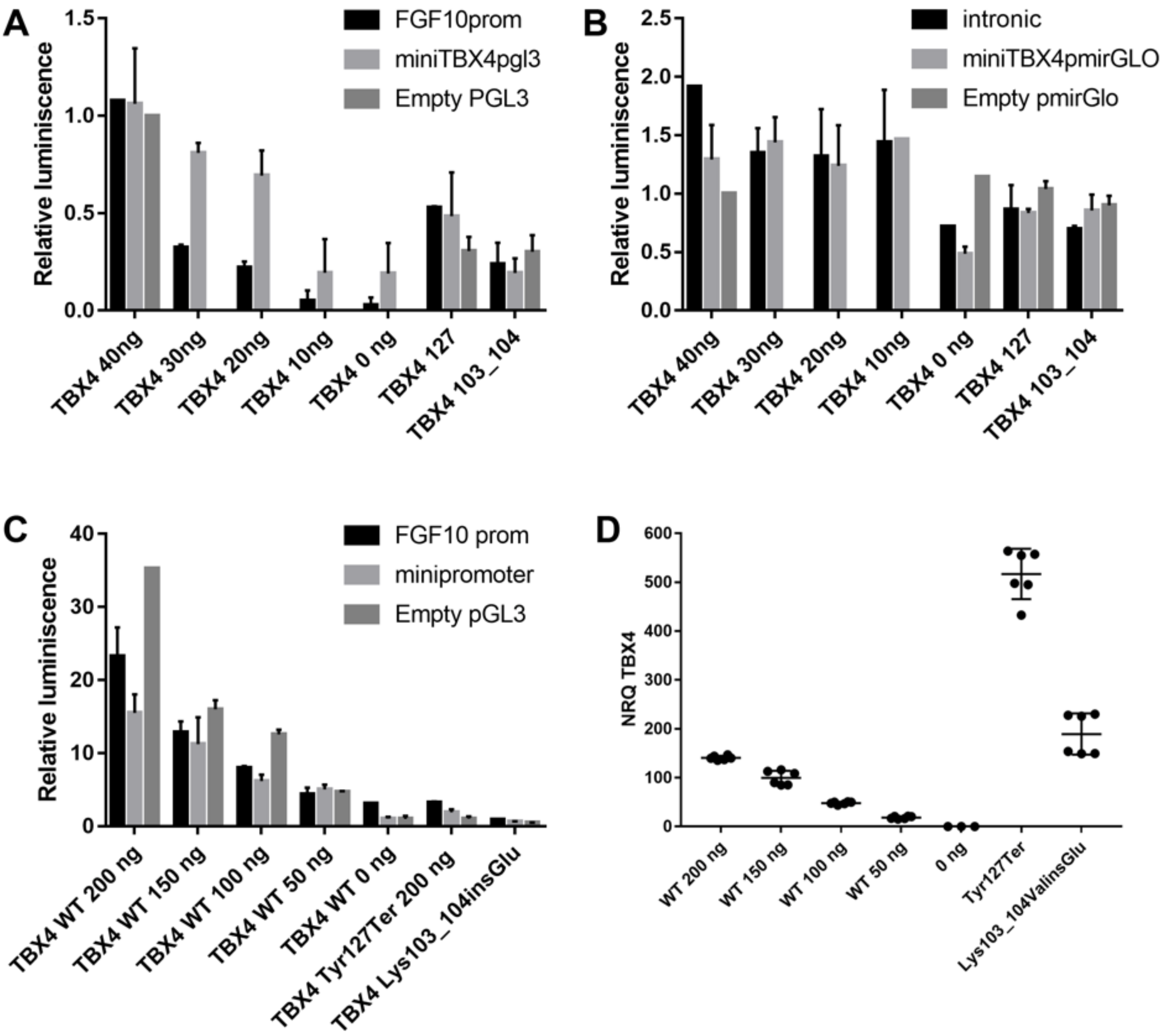


**eFigure 8. Optimization of the luciferase assay.** A) Titration of TBX4 levels in the different constructs in a 96-well plate (FGF10 promoter, mini TBOX and empty pGL3), p.Tyr127Ter and p.Lys103_Val104insGlu were used as pathogenicity controls, 40 ng of each luciferase construct were used for all the conditions. The luciferase activities of each construct increased proportionally to the transfected amount of *TBX4*. B) Titration of TBX4 levels of the different constructs in a 96-well plate (FGF10 intronic island, mini TBOX and empty pGL3), p.Tyr127Ter and

p.Lys103_Val104insGlu were used as pathogenicity controls, 40 ng of each luciferase construct were used for all the conditions. None of the constructs responded to TBX4 in a stable dose-dependent manner. C) Titration of TBX4 levels in a 24-well plate, 200 ng of each luciferase construct were used for all the conditions. D) *TBX4* mRNA levels quantified by qPCR. *TBX4* mRNA levels increased proportionally as the ng used for the transfection, 200 ng of the two pathogenic variants were used. All the data are shown as mean ± SD (n = 2).

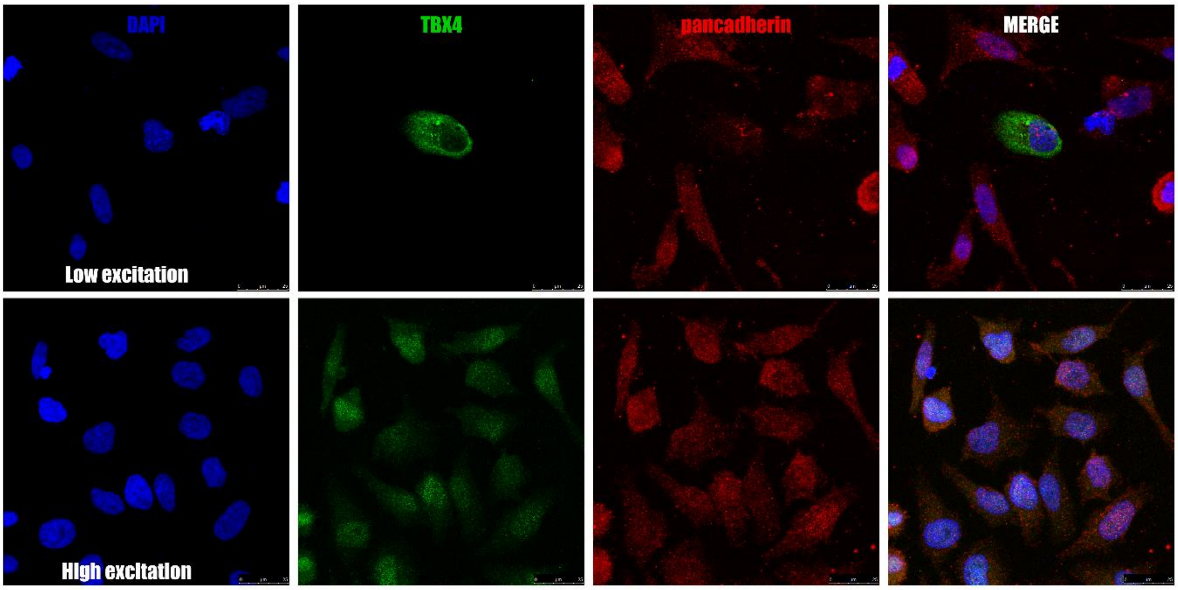


**eFigure 9. Loss of immunoreactivity of the p.Ile270Ser *TBX4* variant.** When we used low excitation settings we were able to detect small apoptotic bodies, while with high excitation we detected only autofluorescence (n=2).

**B.** ***In silico***

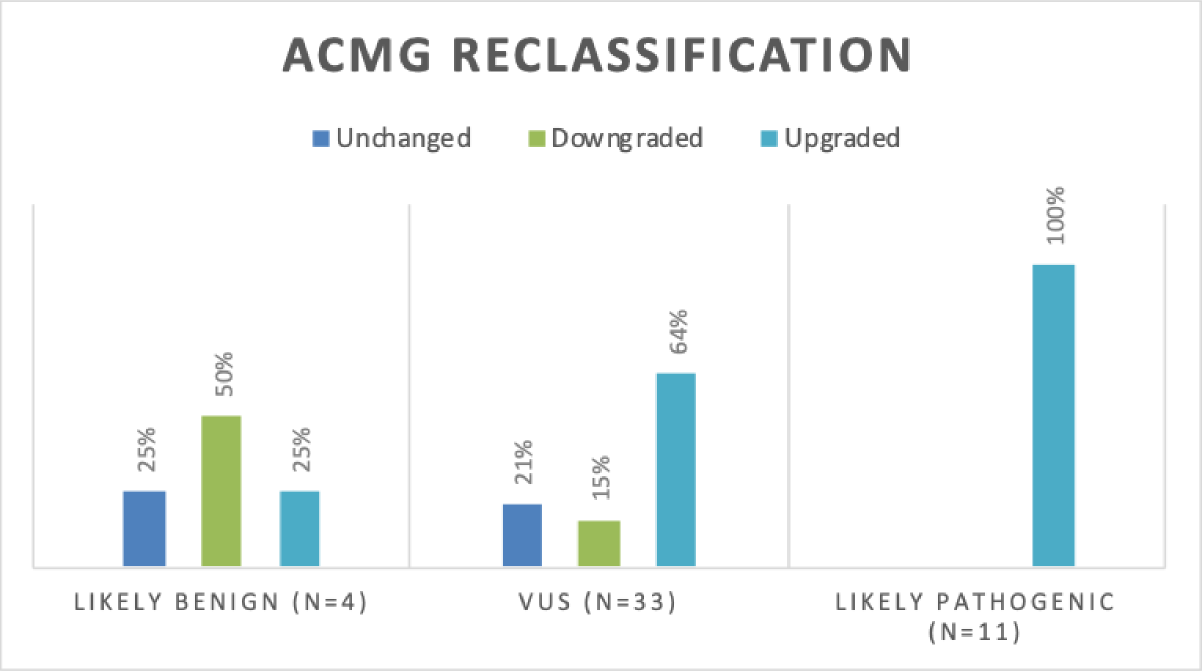


**eFigure 4. Representation of the percentage number of variants with altered classification following functional assessment by luciferase assay.** As per guidelines issued by the American College of Medical Genetics and Genomics (ACMG), interpretation of *TBX4* variants was amended following the application of the functional evidence criterion. Abbreviations: VUS, variant of uncertain significance.

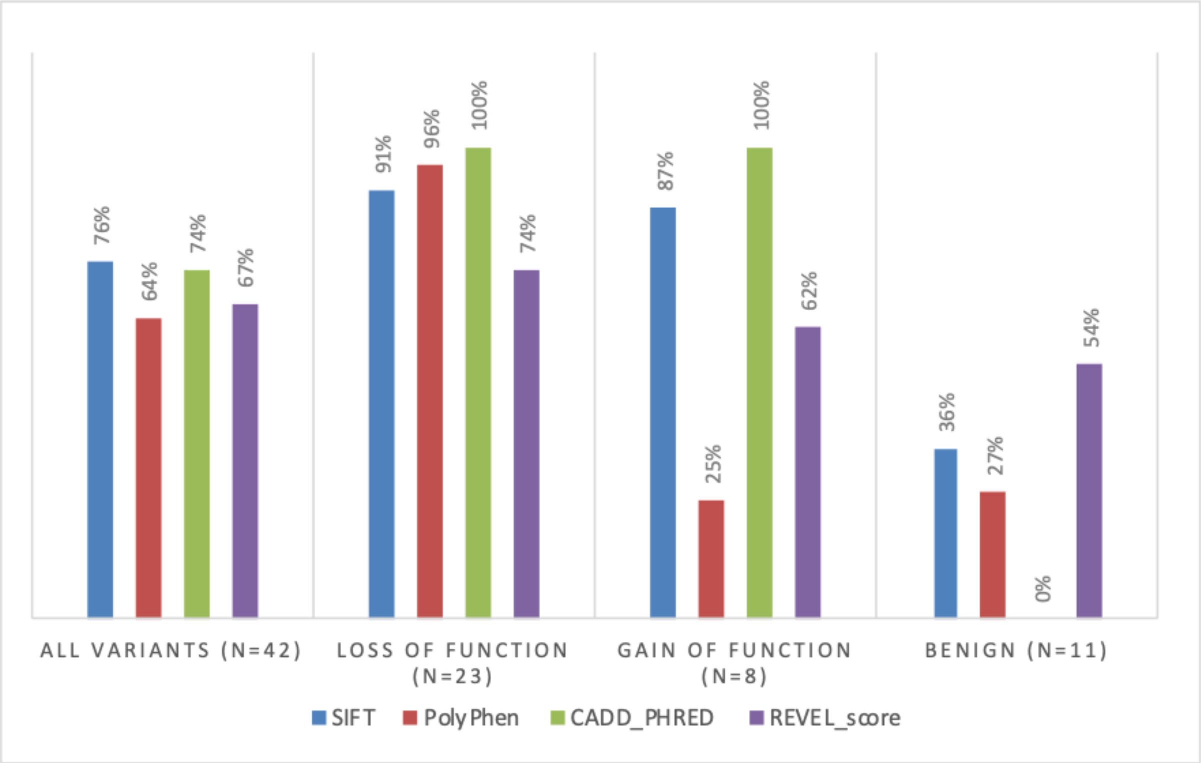


**eFigure 5. Comparison of the accuracy of different *in silico* prediction tools.** Percentage number of functionally assessed *TBX4* variants (by luciferase assay) where pathogenicity was correctly predicted by SIFT (Sorting Intolerant From Toleran) (9), PolyPhen (Polymorphism Phenotyping) (10), CADD (Combined Annotation Dependent Depletion) (11), and REVEL (Rare Exome Variant Ensemble Learner) (12). Overall, the predictors classified correctly similar percentages of variants. When interpreting gain-of-function variants, the CADD score was the most reliable package with noted inconsistencies across different tools. For evidence supportive of pathogenicity, we applied the following criteria: SIFT ≠ tolerated; PolyPhen ≠ benign; CADD ≥ 15; REVEL ≥ 0.5.

#### III. Clinical

##### Radiological substudy

We obtained computed tomography (CT) images of the chest for 13 TBX4 cases recruited to the National Institute for Health Research BioResource–Rare Diseases (NBR) study; all scans were performed at the time of PAH diagnosis. Five of the above individuals were carriers of truncating (frameshift and nonsense) *TBX4* variants. Out of the remaining 8 cases with missense variants, only 3 were classed as likely pathogenic/pathogenic (taking into account our functional work) with the remaining 5 excluded from the radiological substudy (total TBX4 case n = 8). Findings were compared to CT diagnostic scans from PAH cases with *BMPR2* mutations (n = 34) and no mutations (n = 143) analyzed on the open-source software 4 Horos (Annapolis, MD USA) by the same cardiothoracic radiologist (AS), blinded to the underlying diagnosis, smoking status, and genotype. Radiological features were scored semi-quantitatively using a customized proforma (eTable 4).

**eTable 4. Reporting proforma for analysis of radiological features.**

Abbreviations: CTPA - Computerized Tomography Pulmonary Angiogram, HRCT - High-Resolution Computerized Tomography, GGO - ground-glass opacities, PVOD - Pulmonary veno-occlusive disease.

| **Parameter** | **Response** |
|---|---|
| ID | character |
| Reader | character |
| CT scan date | date |
| Slice thickness | numeric |
| Number of slices | numeric |

| | |
|---|---|
| CTPA | done/not done |
| HRCT | done/not done |
| Expiratory CT | done/not done |
| Pleural effusion | Nil;Trace;Mild;Moderate;Severe |
| Subcutaneous oedema | Present, absent |
| Severity of GGO centrilobular pattern | None;Subtle;Present |
| Severity of GGO non-specific mosaic pattern | None;Subtle;Present |
| Distribution of GGO | C-central; U-upper; Z-zonal; D-diffuse |
| Pulmonary arteriovenous malformations | Present, absent |
| Largest bronchial artery size | Numeric (mm) |
| Mediastinal venous collaterals | Present, absent |
| Intralobular septal thickening | None;Subtle;Present |
| Mediastinal lymphadenopathy | None;Subtle;Present |
| Emphysema | None;Subtle;Present |
| Bronchial wall thickening | None;Subtle;Present |
| Fibrosis | None;Subtle;Present |
| Air trapping | None;Subtle;Present |
| Suspected PVOD | Yes, no |

#### Histopathology

Lung histology was available for 17 previously published *TBX4* cases (supplementary data, xlsx). Alongside varying degrees of pulmonary vascular hypertensive changes, parenchymal abnormalities were present in all individuals. Alveolar features ranged from thin septa to diffuse dysplasia representing growth arrest at different stages of lung development. On the severe end of the spectrum, out of 4 cases with lethal lung maldevelopment, 3 had mutations in the T-BOX domain including 2 loss-of-function missense variants affecting the same amino-acid position (p.Glu86Gln and p.Glu86Lys) and 1 protein-truncating variant (c.524_527del). The fourth case carried a paternally inherited *TBX4* missense variant at the C-terminus (c.1198G>A) shown to be benign by our functional analyses; this was an infant who died at 4 months due to progressively worsening PAH with respiratory failure. As reported by the authors, trio exome sequencing (proband and parents) also showed a *de novo* nonsense variant in the *CTNNB1* gene which may account for some of the observed phenotypic features, including microcephaly and muscle spasticity.

Interstitial fibrotic changes were described in 9 individuals including 2 cases with non-specific interstitial pneumonia (NSIP). All of the above were heterozygous for either missense loss-of-function or protein-truncating *TBX4* variants the majority of which (6/9) were located in the T-BOX domain, including the 2 NSIP cases. Bronchial abnormalities of varying degrees were reported in 8/17 individuals. A single patient with histological findings of pulmonary veno-occlusive disease (PVOD) harbored a missense variant (c.1145A>C ) shown to be benign by our functional analyses. Another individual was heterozygous for a pathogenic missense variant (c.432G>T) inducing gain-of-function with reported typical findings of PAH in the explanted lung tissue alongside emphysematous changes.

#### IV. Supplemental References

1. Stauffer W, Sheng H, Lim HN. EzColocalization: An ImageJ plugin for visualizing and measuring colocalization in cells and organisms. *Scientific Reports*

2018;8:15764.

2. Richards S, Aziz N, Bale S, Bick D, Das S, Gastier-Foster J, Grody WW, Hegde M, Lyon E, Spector E, Voelkerding K, Rehm HL. Standards and guidelines for the interpretation of sequence variants: A joint consensus recommendation of the American College of Medical Genetics and Genomics and the Association for Molecular Pathology. *Genetics in Medicine* 2015;17:405–424.
3. Whiffin N, Minikel E, Walsh R, O'Donnell-Luria AH, Karczewski K, Ing AY, Barton PJR, Funke B, Cook SA, MacArthur D, Ware JS. Using high-resolution variant frequencies to empower clinical genome interpretation. *Genet Med* 2017;19:1151–1158.
4. Eyries M, Montani D, Nadaud S, Girerd B, Levy M, Bourdin A, Trésorier R, Chaouat A, Cottin V, Sanfiorenzo C, Prevot G, Reynaud-Gaubert M, Dromer C, Houeijeh A, Nguyen K, Coulet F, Bonnet D, Humbert M, Soubrier F. Widening the landscape of heritable pulmonary hypertension mutations in paediatric and adult cases. *Eur Respir J* 2019;53:.
5. Zhu N, Gonzaga-Jauregui C, Welch CL, Ma L, Qi H, King AK, Krishnan U, Rosenzweig EB, Ivy DD, Austin ED, Hamid R, Nichols WC, Pauciulo MW, Lutz KA, Sawle A, Reid JG, Overton JD, Baras A, Dewey F, Shen Y, Chung WK. Exome Sequencing in Children With Pulmonary Arterial Hypertension Demonstrates Differences Compared With Adults. *Circ Genom Precis Med* 2018;11:e001887.
6. Gräf S, Haimel M, Bleda M, Hadinnapola C, Southgate L, Li W, Hodgson J, Liu B, Salmon RM, Southwood M, Machado RD, Martin JM, Treacy CM, Yates K, Daugherty LC, Shamardina O, Whitehorn D, Holden S, Aldred M, Bogaard HJ, Church C, Coghlan G, Condliffe R, Corris PA, Danesino C, Eyries M, Gall H, Ghio

S, Ghofrani H-AA, *et al.* Identification of rare sequence variation underlying heritable pulmonary arterial hypertension. *Nature Communications* 2018;9:1416.

7. Thoré P, Girerd B, Jaïs X, Savale L, Ghigna M-R, Eyries M, Levy M, Ovaert C, Servettaz A, Guillaumot A, Dauphin C, Chabanne C, Boiffard E, Cottin V, Perros F, Simonneau G, Sitbon O, Soubrier F, Bonnet D, Remy-Jardin M, Chaouat A, Humbert M, Montani D. Phenotype and outcome of pulmonary arterial hypertension patients carrying a TBX4 mutation. *Eur Respir J* 2020;55:1902340.
8. Wickham H. *ggplot2: Elegant Graphics for Data Analysis*, 2nd ed. 2016. Cham: Springer International Publishing : Imprint: Springer; 2016. doi:10.1007/978-3-319-24277-4.
9. Sim NL, Kumar P, Hu J, Henikoff S, Schneider G, Ng PC. SIFT web server: Predicting effects of amino acid substitutions on proteins. *Nucleic Acids Research* 2012;40:W452–W457.
10. Adzhubei I, Jordan DM, Sunyaev SR. Predicting functional effect of human missense mutations using PolyPhen-2. *Current Protocols in Human Genetics* 2013;Chapter 7:Unit7.20.
11. Rentzsch P, Witten D, Cooper GM, Shendure J, Kircher M. CADD: predicting the deleteriousness of variants throughout the human genome. *Nucleic Acids Research* 2018;doi:10.1093/nar/gky1016.
12. Ioannidis NM, Rothstein JH, Pejaver V, Middha S, McDonnell SK, Baheti S, Musolf A, Li Q, Holzinger E, Karyadi D, Cannon-Albright LA, Teerlink CC, Stanford JL, Isaacs WB, Xu J, Cooney KA, Lange EM, Schleutker J, Carpten JD, Powell IJ, Cussenot O, Cancel-Tassin G, Giles GG, MacInnis RJ, Maier C, Hsieh C-L, Wiklund F, Catalona WJ, Foulkes WD, *et al.* REVEL: An Ensemble Method for Predicting the Pathogenicity of Rare Missense Variants. *American Journal of*

*Human Genetics* 2016;99:877.
